# Characterising the contribution of rare protein-coding germline variants to prostate cancer risk and severity in 37,184 cases

**DOI:** 10.1101/2024.05.10.24307164

**Authors:** Jonathan Mitchell, Niedzica Camacho, Patrick Shea, Konrad H. Stopsack, Vijai Joseph, Oliver Burren, Ryan Dhindsa, Abhishek Nag, Jacob E. Berchuck, Amanda O’Neill, Ali Abbasi, Anthony W. Zoghbi, Jesus Alegre-Díaz, Pablo Kuri-Morales, Jaime Berumen, Roberto Tapia-Conyer, Jonathan Emberson, Jason M. Torres, Rory Collins, Quanli Wang, David Goldstein, Athena Matakidou, Carolina Haefliger, Lauren Anderson-Dring, Ruth March, Vaidehi Jobanputra, Brian Dougherty, Keren Carss, Slavé Petrovski, Philip W. Kantoff, Kenneth Offit, Lorelei A. Mucci, Mark Pomerantz, Margarete A. Fabre

## Abstract

The etiology of prostate cancer, the second most common cancer in men globally, has a strong heritable component. While rare coding germline variants in several genes have been identified as risk factors from candidate gene and linkage studies, the exome-wide spectrum of causal rare variants remains to be fully explored. To more comprehensively address their contribution, we analysed data from 37,184 prostate cancer cases and 331,329 male controls from five cohorts with germline exome/genome sequencing and one cohort with imputed array data from a population enriched in low-frequency deleterious variants. Our gene-level collapsing analysis revealed that rare damaging variants in *SAMHD1* as well as genes in the DNA damage response pathway (*BRCA2*, *ATM* and *CHEK2*) are associated with the risk of overall prostate cancer. We also found that rare damaging variants in *AOX1* and *BRCA2* were associated with increased severity of prostate cancer in a case-only analysis of aggressive versus non-aggressive prostate cancer. At the single-variant level, we found rare non-synonymous variants in three genes (*HOXB13*, *CHEK2*, *BIK*) significantly associated with increased risk of overall prostate cancer and in four genes (*ANO7*, *SPDL1*, *AR*, *TERT*) with decreased risk. Altogether, this study provides deeper insights into the genetic architecture and biological basis of prostate cancer risk and severity.

## Introduction

Prostate cancer is the second most common cancer in men globally, with over 1.5 million new cases and 397,000 deaths estimated in 2022^1,2^. Whilst the vast majority of men diagnosed with localised disease are either cured or survive their cancer for many years, the 5-year survival in metastatic cases is just 30%, and a substantial number live with treatment-related morbidity^3,4^.

The pathogenesis of prostate cancer involves complex interactions between inherited genetic features, acquired somatic mutations, and environmental factors. The important role of the germline genome is evident by the high heritability of prostate cancer risk, estimated by twin studies at 57%^5^. While genome-wide association studies (GWAS) have identified 451 risk variants to date, a large proportion of the heritability remains unaccounted for^6–8^. Importantly, emerging evidence suggests that the set of genes influencing the risk of developing prostate cancer is, at least in part, distinct from the genes influencing prostate cancer survival^9^; for example, while a polygenic risk score incorporating disease risk variants increased the risk of both overall and aggressive prostate cancer, it discriminated only modestly between aggressive and non-aggressive prostate cancer^8^.

Rare protein-coding germline variants associated with disease risk and progression, compared to common variants, have larger effect sizes and often directly implicate causal genes^10^, making rare variant disease associations particularly valuable for understanding disease pathogenesis and, as a result, identifying drug targets and elucidating treatment response^11,12^. For prostate cancer, linkage and candidate gene studies have identified influential rare variants in a small number of specific genes, such as *HOXB13* and *BRCA2*^13,14^. To more comprehensively assess the contribution of rare germline variants exome-wide to the development of prostate cancer and its severity, we first tested for rare variant associations at the gene level, utilising global biobanks, curated disease cohorts and clinical trial participants with germline whole exome sequencing (WES) or whole genome sequencing (WGS) data (total 19,926 cases; 187,705 controls), and subsequently at the single variant level, additionally incorporating imputed array data from the FinnGen cohort^15^ (total 33,608 cases; 309,439 controls). The scale of meta-analysis presented here represents the most comprehensive characterisation to date of the role rare coding germline variants play in prostate cancer pathogenesis.

## Results

### Gene-level association testing

To investigate the aggregated influence of rare germline variants on prostate cancer risk and severity at the level of individual genes, we meta-analysed WES and WGS data from five cohorts totalling 19,926 prostate cancer cases and 187,705 male controls that met all quality control criteria (Table 1, see Methods). These cohorts comprised The UK Biobank (UKB)^16,17^, The Mexico City Prospective Study (MCPS)^18,19^, The 100,000 Genomes Project (100KGP)^20,21^, three studies within the New York-Boston-AstraZeneca (NYBAZ) prostate cancer study, and a collection of AstraZeneca clinical trial (AZCT) participants. Except MCPS, which predominantly comprises individuals with Admixed American ancestry, the cohorts are primarily of European ancestry. However, we additionally included African, East Asian and South Asian strata where sufficient numbers of individuals were available.

**Table 1:**
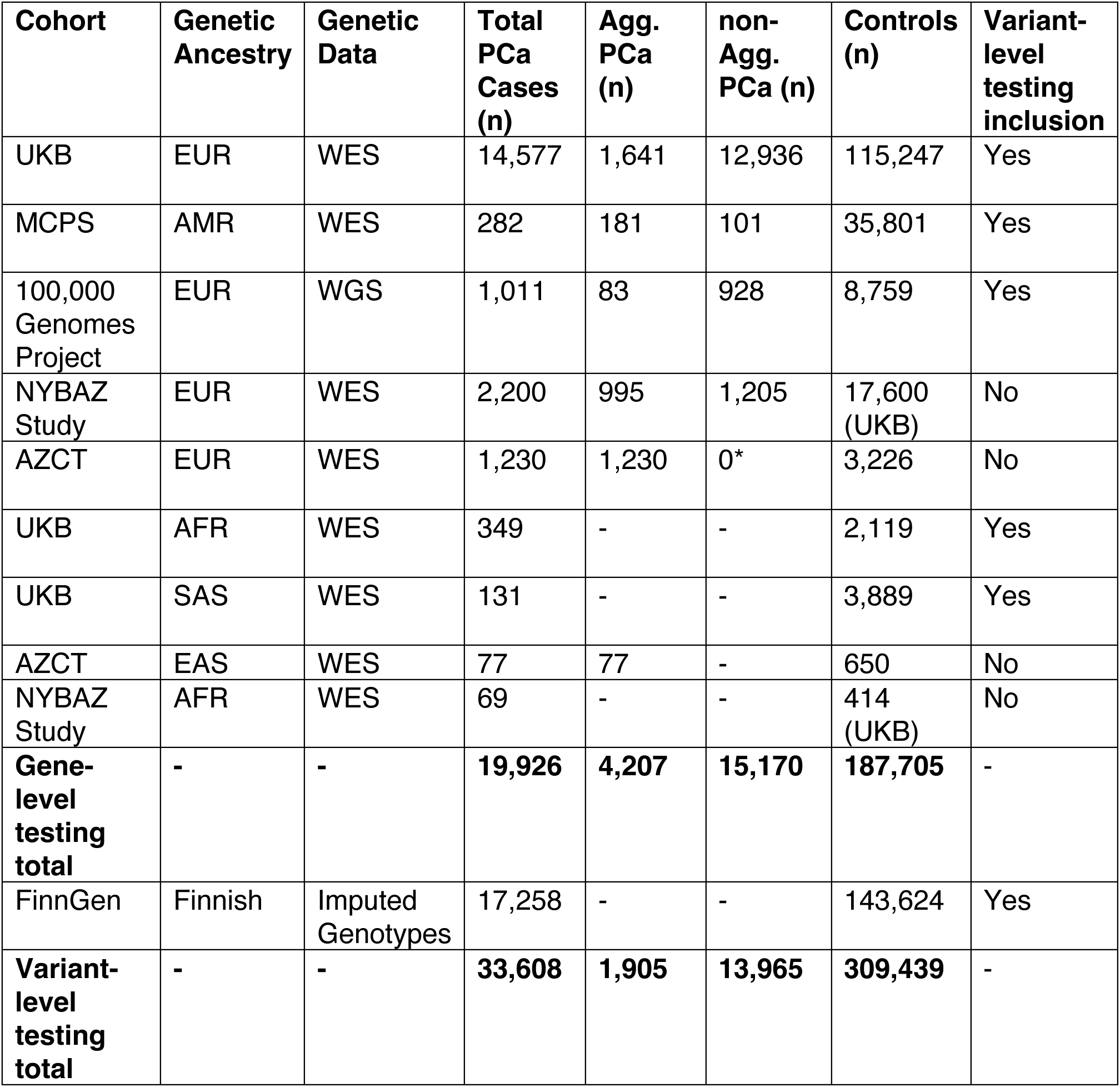
Sample size, ancestry and genetic data type of all cohorts used in the gene-level and single variant-level genetic association meta-analyses. Aggressive prostate cancer (agg. PCa) is defined by tumour stage T4/N1/M1, Gleason score >=8, prostate cancer as underlying cause of death, metastatic prostate cancer, prostate cancer treated with chemotherapy or castration resistant prostate cancer. *For the clinical trial cohort agg. PCa Vs non-agg. PCa analysis in EUR, a subset of non-aggressive UKB cases were used as this cohort contained none.

| **Cohort** | **Genetic Ancestry** | **Genetic Data** | **Total PCa Cases (n)** | **Agg. PCa (n)** | **non-Agg. PCa (n)** | **Controls (n)** | **Variant-level testing inclusion** |
| --- | --- | --- | --- | --- | --- | --- | --- |
| UKB | EUR | WES | 14,577 | 1,641 | 12,936 | 115,247 | Yes |
| MCPS | AMR | WES | 282 | 181 | 101 | 35,801 | Yes |
| 100,000 Genomes Project | EUR | WGS | 1,011 | 83 | 928 | 8,759 | Yes |
| NYBAZ Study | EUR | WES | 2,200 | 995 | 1,205 | 17,600 (UKB) | No |
| AZCT | EUR | WES | 1,230 | 1,230 | 0* | 3,226 | No |
| UKB | AFR | WES | 349 | - | - | 2,119 | Yes |
| UKB | SAS | WES | 131 | - | - | 3,889 | Yes |
| AZCT | EAS | WES | 77 | 77 | - | 650 | No |
| NYBAZ Study | AFR | WES | 69 | - | - | 414 (UKB) | No |
| **Gene-level testing total** | - | - | **19,926** | **4,207** | **15,170** | **187,705** | - |
| FinnGen | Finnish | Imputed Genotypes | 17,258 | - | - | 143,624 | Yes |
| **Variant-level testing total** | - | - | **33,608** | **1,905** | **13,965** | **309,439** | - |

Gene-phenotype association testing was performed under the previously described collapsing analysis framework^22,23^. To maximise discovery across potential genetic architectures, we included eleven qualifying variant (QV) models for each gene (ten dominant and one recessive), which filtered variants on a range of predicted effects and population frequency thresholds (Supplementary Table 1). The threshold for a suggestive association was set at *P* < 2.6×10^-6^ (Bonferroni corrected for the number of genes tested, 0.05 / 18,948 genes), and the study-wide significant threshold at the more stringent *P* < 1×10^-8^, which we have previously shown to result in an extremely low false positive rate when testing multiple QV models across multiple traits^23^.

We first tested for genes associated with the risk of developing prostate cancer overall in a case-control analysis (19,926 cases vs 187,705 controls). The approach was robust, with no significant inflation in test statistics across the eleven QV models (λ_mean_ = 1.04 ± 0.024, Supplementary Figure 1 and Supplementary Table 2). We identified rare protein-truncating variants in the DDR genes *BRCA2* (OR = 3.23 [2.65-3.90], *P* = 7.5×10^-29^) and *ATM* (OR = 2.92 [2.34-3.63], *P* = 1.17×10^-19^), and additionally rare damaging variants in *SAMHD1* (OR = 2.02 [1.65-2.45], *P* = 2.36×10^-11^) as significantly associated with increased prostate cancer risk (Figures 1 & 2, Table 2 and Supplementary Table 3). Germline variants in *SAMHD1* have recently been reported as associated with the risk of prostate cancer in an analysis of the UK Biobank only^24^, and additionally, there is evidence of association with breast cancer^25^ and primary cancers collectively^26,27^. Here, we independently validate the *SAMHD1* association with prostate cancer in the UK Biobank and replicate the finding in additional cohorts (Supplementary Table 3). Rare damaging variants in *CHEK2* (OR = 1.69 [1.41-2.01], *P* = 2.69×10^-8^) and rare synonymous variants in *DMD* (OR = 0.50 [0.36-0.67], *P* = 8.6×10^-7^) were associated with prostate cancer risk at the suggestive significance threshold. Although the Duchenne muscular dystrophy (DMD) gene has been previously implicated in cancer^28^, the association we report here is for synonymous variants and at the suggestive level, and should therefore be interpreted with caution. *TET2* was also significantly associated with prostate cancer risk (OR = 3.31 [2.26-4.78], *P* = 1.71×10^-9^). However, the strong correlation between *TET2* carrier status and age (UKB EUR cohort: *P* = 3.25×10^-5^), and the skewed distribution of alternate reads percentage to below 50% (Supplementary Figure 2), indicates a somatic mutational process. Indeed, while our analysis is confounded by age, the causal association of clonal somatic variants in the well-established clonal haematopoiesis (CH) driver gene *TET2* and prostate cancer has been described previously^29^.

**Figure 1:**
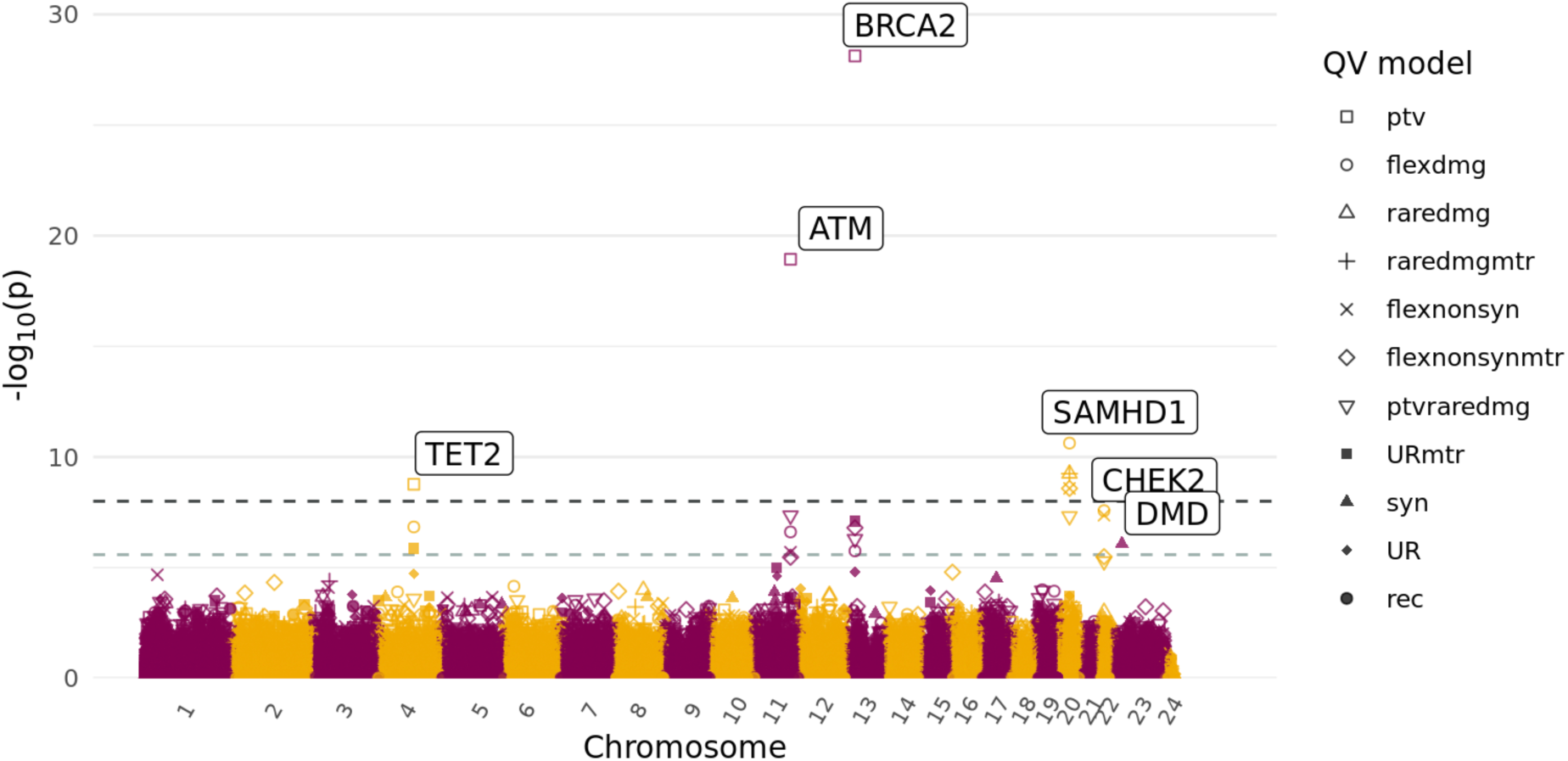
Manhattan plot of all meta-analysis gene-level association tests with risk of developing overall prostate cancer. The x-axis is the genomic position of the gene, and the y-axis is the -log_10_ transformed unadjusted *P*-values for all qualifying variant models as indicated in the legend. The light grey dashed line represents the suggestive significance threshold (*P* = 2.6×10^-6^) and the dark grey dashed line the study-wide significance threshold (*P* = 1×10^-8^). Genes which reach the suggestive significance threshold are labelled, and only the most significant qualifying variant model for each gene is labelled. Qualifying variant models defined in Supplementary Table 1.

**Figure 2:**
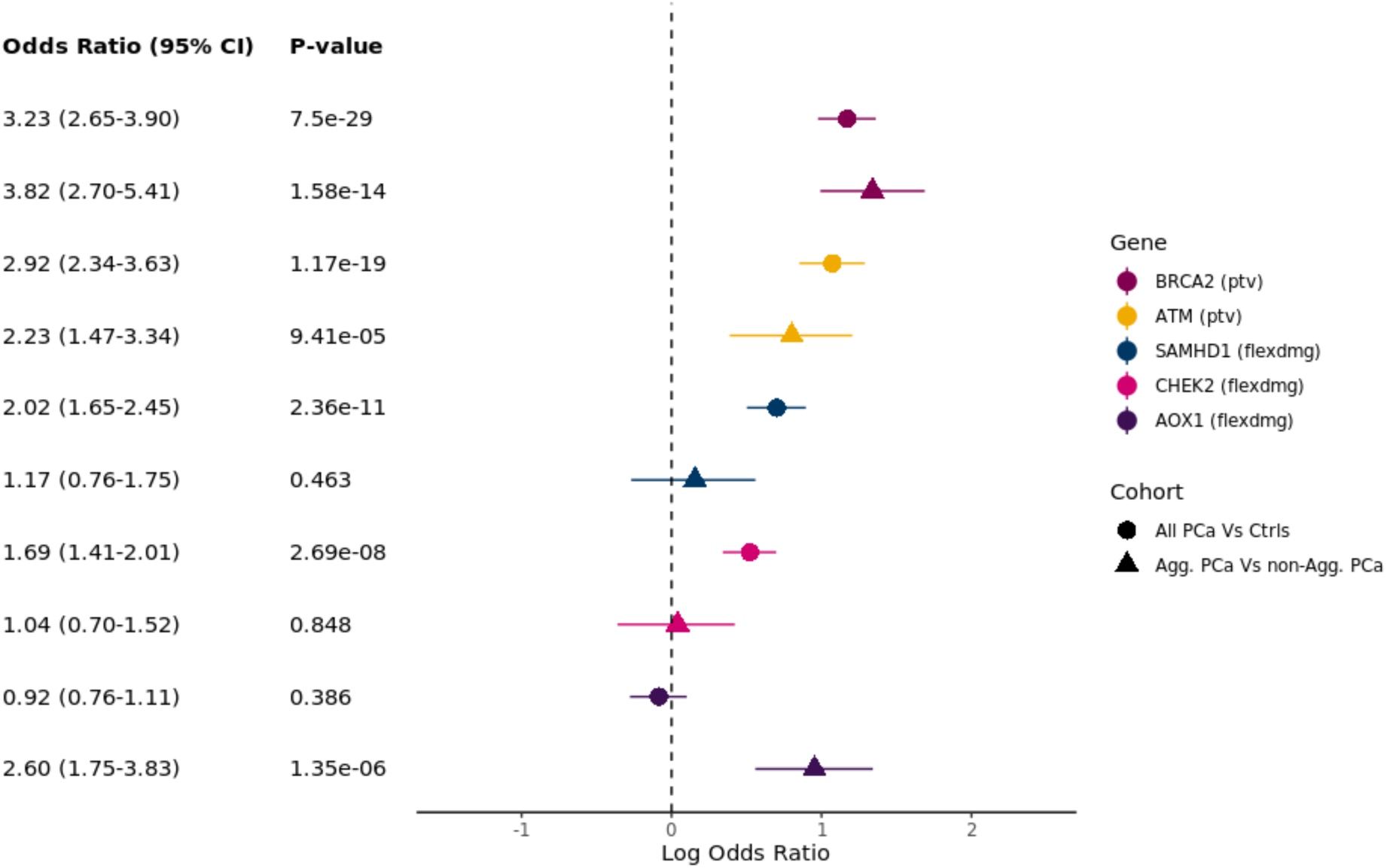
Forest plot showing the association of genes with prostate cancer risk (All PCa Vs Ctrls) and severity (Agg. PCa Vs Ctrls) which reached the suggestive significance threshold (*P* = 2.6×10^-6^) for non-synonymous qualifying variant (QV) models. Gene and QV model are as indicated in legend. For genes where more than one QV model (defined in Supplementary Table 1) passed the suggestive significance threshold the most significant is plotted. ptv = rare protein truncating variant QV model; flexdmg = rare damaging non-synonymous QV model. PCa = prostate cancer; Agg = aggressive. Non-agg = non-aggressive.

**Table 2:** Genes significantly associated at the suggestive significance level (*P* < 2.6×10^-6^) with risk of developing prostate cancer (PCa Vs Ctrls) and its severity (Agg. PCa Vs non-Agg. PCa). The qualifying variant model (defined in Supplementary Table 1) with the strongest association is shown. Carrier frequency is the fraction of individuals with at least one qualifying allele in the gene.

| Gene | QV Model | Association Analysis | P | OR [95% CI] | Case Carrier Frq. | Ctrl Carrier Frq. |
| --- | --- | --- | --- | --- | --- | --- |
| *BRCA2* | ptv | PCa Vs Ctrls | 7.50x<br>10<sup>-29</sup> | 3.23<br>[2.65-3.90] | 0.00793 | 0.00248 |
|  |  | Agg. PCa Vs non-Agg. PCa | 1.58x<br>10<sup>-14</sup> | 3.82<br>[2.70-5.41] | 0.0194 | 0.00492 |
|  |  | Agg. PCa Vs Ctrls | 1.47x10<sup>-36</sup> | 8.23 [6.17-10.85] | 0.0188 | 0.00246 |
| *ATM* | ptv | PCa Vs Ctrls | 1.17x<br>10<sup>-19</sup> | 2.92<br>[2.34-3.63] | 0.00607 | 0.00225 |
|  |  | Agg. PCa Vs non-Agg. PCa | 9.41x<br>10<sup>-05</sup> | 2.23<br>[1.47-3.34] | 0.0111 | 0.00478 |
|  |  | Agg. PCa Vs Ctrls | 1.74x10<sup>-16</sup> | 5.27 [3.65-7.46] | 0.0116 | 0.00221 |
| *SAMHD1* | flexdmg | PCa Vs Ctrls | 2.36x<br>10<sup>-11</sup> | 2.02<br>[1.65-2.45] | 0.00678 | 0.00322 |
|  |  | Agg. PCa Vs non-Agg. PCa | 0.463 | 1.17<br>[0.76-1.75] | 0.00823 | 0.00810 |
|  |  | Agg. PCa Vs Ctrls | 5.98x10<sup>-3</sup> | 1.80 [1.17-2.68] | 0.00737 | 0.00341 |
| *CHEK2* | flexdmg | PCa Vs Ctrls | 2.69x<br>10<sup>-8</sup> | 1.69<br>[1.41-2.01] | 0.00793 | 0.00522 |
|  |  | Agg. PCa Vs non-Agg. PCa | 0.848 | 1.04<br>[0.70-1.52] | 0.00944 | 0.00904 |
|  |  | Agg. PCa Vs Ctrls | 8.04x10<sup>-3</sup> | 1.65 [1.13-2.34] | 0.00951 | 0.00510 |
| *AOX1* | flexdmg | PCa Vs Ctrls | 0.386 | 0.92<br>[0.76-1.11] | 0.00642 | 0.00889 |
|  |  | Agg. PCa Vs non-Agg. PCa | 1.35x<br>10<sup>-6</sup> | 2.60<br>[1.75-3.83] | 0.0128 | 0.00485 |
|  |  | Agg. PCa Vs Ctrls | 6.56x10<sup>-3</sup> | 1.54 [1.12-2.09] | 0.0121 | 0.00870 |

Consistent with the known importance of the DNA damage response (DDR) pathway in prostate cancer pathogenesis^14^, we found *BRCA2*, *ATM* and *CHEK2* to be among the most significant risk genes. In the UK Biobank 267/14,577 (1.8%) individuals who developed prostate cancer carried a qualifying variant in one of these three genes, compared to 900/115247 (0.8%) controls (*P* = 1.12×10^-29^). Several other DDR genes, with a lower frequency of qualifying variant carriers, demonstrated association with the risk of prostate cancer development at a nominal significance level, including *MSH2* (OR = 3.38 [1.55-6.90], *P* = 1.20×10^-3^) and *NBN* (OR = 1.92 [1.22-2.93], *P* = 3.35×10^-3^, Supplementary Figure 3 and Supplementary Table 4).

Next, using the available clinical data for the five cohorts, we stratified cases into aggressive prostate cancer (agg. PCa) and non-aggressive prostate cancer (non-agg. PCa), a distinction we refer to subsequently as ‘severity’. Aggressive prostate cancer was defined if any one of a number of criteria were met: tumour stage T4 or N1 or M1, Gleason score >=8, prostate cancer as primary cause of death, prostate cancer treated with chemotherapy, or castration-resistant prostate cancer (see Methods). We performed a case-only gene-level association test across the exome (4,207 agg. PCa cases vs 15,170 non-agg. PCa cases, Table 1), to identify genes associated with disease severity (Table 2, Supplementary Tables 5 & 6 and Supplementary Figures 4 & 5). Protein-truncating variants in *BRCA2* (OR = 3.82 [2.70-5.41], *P* = 1.58×10^-14^) were significantly associated with increased severity, as were rare damaging variants in *AOX1* at the suggestive level (OR = 2.60 [1.75-3.83], *P* = 1.35×10^-6^). Consistent with our association of rare variants in *AOX1* specifically with prostate cancer severity, a prior GWAS identified a common variant at the *AOX1* locus – that correlated with *AOX1* expression levels – associated with prostate-cancer-specific survival time, and that expression levels of *AOX1* were associated with biochemical recurrence of prostate cancer^30^. Additionally, of the genes significantly associated with the risk of developing prostate cancer, the DDR gene *ATM* was nominally associated with severity (OR = 2.23 [1.47-3.34], *P* = 9.41×10^-5^, Supplementary Table 7 and Supplementary Figure 6). Indeed, we found that in UKB 48/1641 (2.9%) of all aggressive prostate cancer cases carried a *BRCA2* or *ATM* qualifying variant compared to 114/12,936 (0.9%) of non-aggressive prostate cancer cases (*P* = 1.46×10^-10^).

Finally at the gene-level, we tested for association between aggressive prostate cancer and controls, and found that protein truncating variants in *BRCA2* (OR = 8.23 [6.17-10.85], *P* = 1.47×10^-36^) and *ATM* (OR = 5.27 [3.66-7.46], *P* = 1.74×10^-16^) were significantly associated in this analysis (Supplementary Tables 8, 9 & 10, Supplementary Figures 7, 8 & 9). Consistent with *BRCA2* and *ATM* being associated with severity, their effect sizes were larger in this aggressive prostate cancer versus controls analysis compared to the overall prostate cancer versus controls analysis (*BRCA2* OR = 3.23 [2.65-3.90] and *P* = 7.5×10^-29^; *ATM* OR = 2.92 [2.34-3.63] and *P* = 1.17×10^-19^).

For three of the genes associated with the risk of developing prostate cancer (*CHEK2*, *SAMHD1*) and prostate cancer severity (*AOX1*) the most significant QV model consisted of a combination of rare predicted damaging missense and protein truncating variants. In all three cases there was a nominal association with the QV model containing only truncating variants (*CHEK2*, OR = 1.58 [1.00-2.41], *P* = 0.035; *SAMHD1*, OR = 2.15 [1.22-3.63], *P* = 0.006; *AOX1*, OR = 3.63 [1.36-9.63], *P* = 0.006) strongly suggesting that the association of these three genes with prostate cancer is through a loss-of-function mechanism. In all genes found to be associated with prostate cancer, the most significantly associated QV model identified a large number of separate qualifying variants distributed along the amino acid sequence (Supplementary Figures 10 & 11).

### Protein-coding variant-level association testing

We next performed a variant-level, exome-wide association study (ExWAS) to identify individual rare variants associated with prostate cancer. We analysed sequencing data from the cohorts included in the gene-level analysis and imputed genotype array data from the FinnGen cohort^15^. The meta-analysis was restricted to cohorts with a low level of genomic inflation within the ExWAS (Supplementary Table 11, see Methods), resulting in a total of 33,608 prostate cancer cases and 309,439 male controls, and we set a threshold of *P* < 1×10^-8^ for study-wide statistical significance^23^.

We identified 92 variants associated with the risk of developing prostate cancer at the study-wide significance threshold, of which sixteen were rare (MAF<1%) in non-Finnish Europeans (Figure 3, Supplementary Table 12). These sixteen rare protein-coding variants were spread over eight loci, and there was statistical evidence for seven of them being the causal variant in the locus (FinnGen SuSiE^31^ posterior inclusion probability (PIP) > 0.05, Table 3). One of the sixteen variants (17:47809406:G:A in *OSBPL7*) was not present in FinnGen and instead associated in the UKB ExWAS (*P* = 2.52×10^-11^), but was not significant (*P* = 0.12) after conditioning on the lead variant (17:48728343:C:T in *HOXB13*) in the locus. All seven putatively causal variants were non-synonymous: a frameshift variant in *CHEK2* (OR = 1.67 [1.46-1.91], P = 1.17×10^-20^), missense variants in *HOXB13* (OR = 4.69 [4.22-5.21], *P* = 1.95×10^-181^), *ANO7* (OR = 0.70 [0.66-0.74], *P* = 4.57×10^-26^), *SPDL1* (OR = 0.72 [0.66-0.78], *P* = 3.06×10^-13^), *AR* (OR = 0.71 [0.66-0.76], *P* = 1.28×10^-11^) and *TERT* (OR = 0.13 [0.07-0.25], *P* = 4.67×10^-10^), and a conservative inframe deletion in *BIK* (OR = 2.08 [1.68-2.57], *P* = 2.04×10^-11^). In the case-only and case-control analyses of aggressive prostate cancer, which were limited to the UKB, MCPS and 100,000 Genomes Project cohorts, there were no significantly associated rare variants.

**Figure 3:**
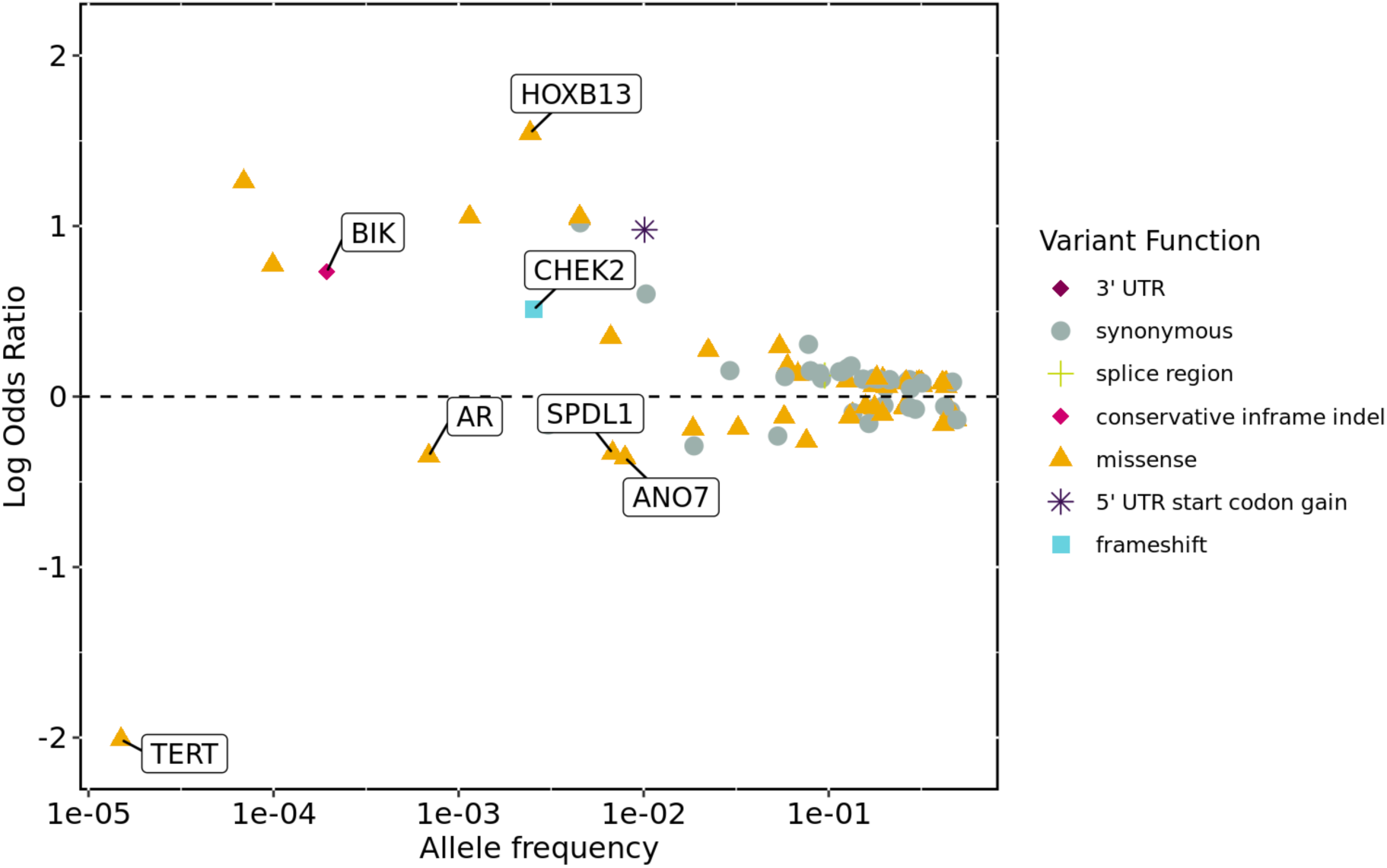
Summary of ExWAS variants which reached study-wide significance (*P*<1×10^-8^) in the meta-analysis for the risk of developing prostate cancer. The x-axis is the variant minor allele frequency in non-Finnish Europeans, and the y-axis is the variant effect estimate. Gene labelled variants are those which are rare in non-Finnish Europeans (Allele Frequency < 1%) and had a posterior inclusion probability of being a causal variant greater than 0.05 in the FinnGen study. The *P*-value used to determine significance is from the Stouffer’s meta-analysis and as this does not generate an effect-size we report here the effect estimate from the FinnGen cohort.

**Table 3:**
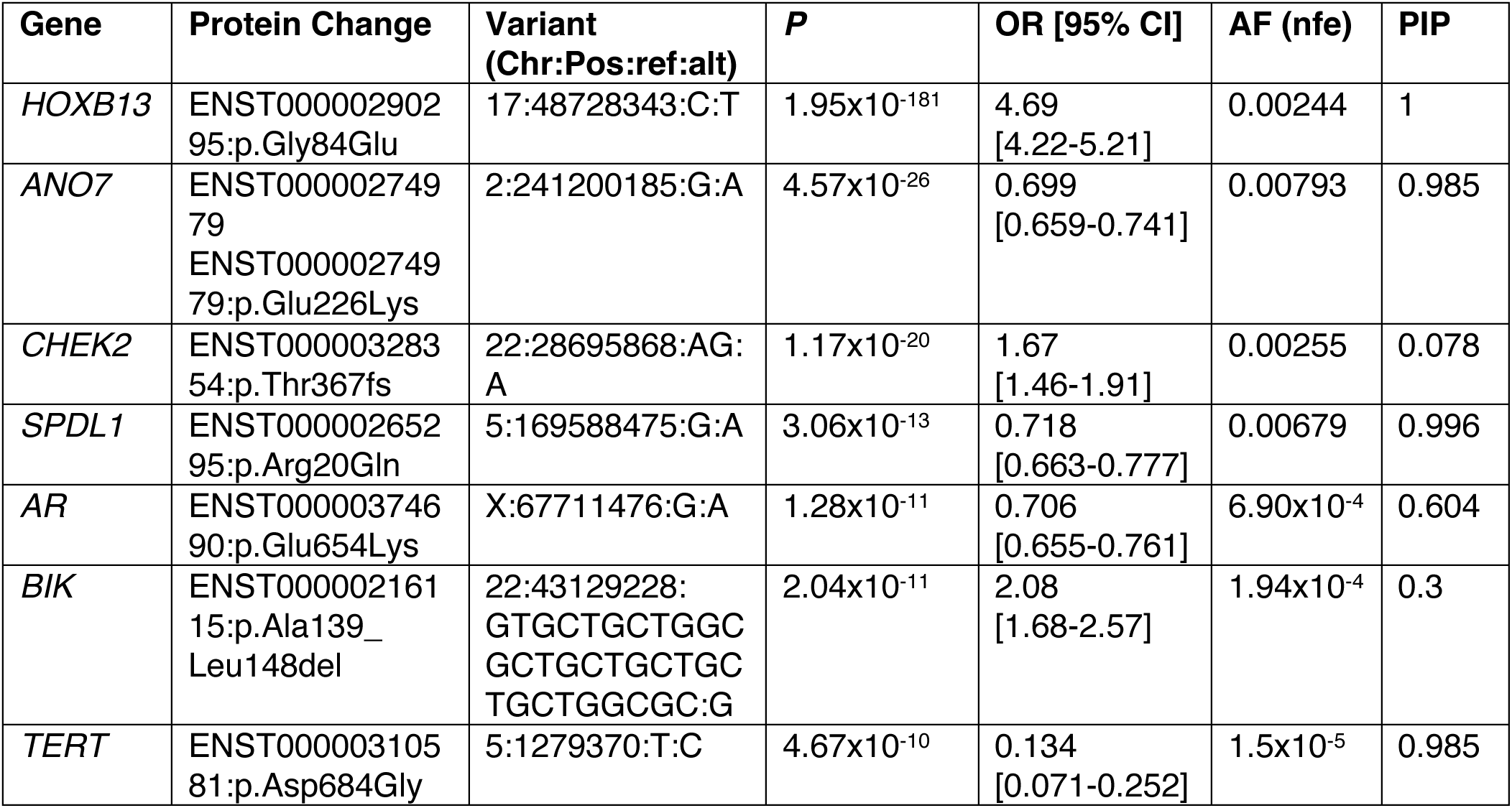
Rare putatively causal variants significantly associated at the study-wide level (*P* < 1×10^-8^) with the risk of prostate cancer. The *P*-value reported is from the Stouffer’s meta-analysis and as this does not generate an effect-size we report here the effect estimate (OR) from the FinnGen cohort. The allele frequency (AF) is taken from the gnomAD non-Finnish European (nfe) population. Posterior inclusion probability (PIP) as calculated by SuSiE in FinnGen.

The rare missense variant in *HOXB13* we identified is the established p.Gly84Glu (17:48728343:C:T) prostate cancer risk variant^13^. Two features are notable. First is the magnitude of risk conferred by the presence of this variant, with carriers in the current study having a more than 4.5 greater odds of developing prostate cancer compared to non-carriers, a level of risk in excess of that imparted by *BRCA2* germline pathogenic variants in most studies^32^. Secondly, our observation that the rare germline *HOXB13* variant is strongly associated with disease risk but not with severity (OR = 0.87 [0.48-1.47] in case-only analysis, *P* = 0.70), is consistent with previous reports implicating *HOXB13* in disease initiation but not progression^33^.

The non-synonymous rare variants we identified in *ANO7*, *CHEK2*, *SPDL1*, *AR* and *BIK* have been previously reported in a study of the Finnish population as being associated with prostate cancer risk^15^. *ANO7* is a prostate-specific gene, and consistent with the protective missense variant reported here (2:241200185:G:A, PIP = 0.985), an *ANO7* eQTL (2:241195850:G:A) common in the European population (MAF = 2.10%) has previously been found to be associated with both prostate cancer risk and severity^34^. *SPDL1* is involved in mitotic checkpoint signalling during cell division^35^, and the *SPDL1* missense variant (5:169588475:G:A) protective for prostate cancer in our analysis has previously been shown to increase the risk of idiopathic pulmonary fibrosis (IPF)^36^, consistent with existing literature on shared genetic alterations between cancer and IPF^37^. *AR* encodes the androgen receptor, a critical regulator of prostate cancer pathogenesis. Disruption of androgen signalling is a common therapeutic strategy in prostate cancer, and, in this context, it is notable that humans with the rare missense *AR* variant (X:67711476:G:A) are protected against prostate cancer. While the *BIK* conservative inframe deletion variant was unique to the FinnGen cohort, a separate rare disruptive inframe deletion at the same genomic position was nominally associated with prostate cancer risk in UKB (22:43129228:GTGCTGCTGGCGCTGCTGC:G, OR = 1.49 [1.20-1.85], *P* = 6.20×10^-4^).

Finally, we identified a previously unreported missense variant (5:1279370:T:C, Table 3) in *TERT* (Telomerase reverse transcriptase). *TERT* is a key determinant of the enzymatic activity of telomerase, whose function is critical for telomere maintenance. Links between telomere biology and cancer have been widely reported^38–40^. The disruptive missense variant in *TERT* we discovered to be protective against prostate cancer was particularly striking in the degree of protection conferred; carriers had 86% lower odds of developing prostate cancer compared to non-carriers.

## Discussion

These results, facilitated by the largest meta-analysis of sequenced cohorts performed to date, demonstrate the power of rare germline variation to deepen our understanding of prostate cancer pathogenesis. Analysing associations between germline variation and different disease end-points provides insight into the distinct pathogenic roles of individual genes^9^. Specifically, in our study we identify germline variants in the case versus control analysis that play a role in the overall risk of prostate cancer. Independently, the genetic variants implicated in the within-case aggressive versus non-aggressive analysis play a role in prostate cancer severity.

While several DDR genes, most prominently *BRCA2*, are established as conferring prostate cancer risk and are included in clinical guidelines for germline genetic testing^41^, the significance of genes beyond *BRCA2* is not well understood^14^. Here, we validate *BRCA2*, *ATM* and *CHEK2* deleterious rare variants as significant risk factors for the development of prostate cancer overall, but do not find significant associations with any additional DDR genes after adjusting for multiple testing. Beyond the DDR, we identify rare non-synonymous variants in three genes associated with increased risk of prostate cancer (*HOXB13*, *SAMHD1*, *BIK*) and four with decreased risk (*ANO7*, *SPDL1*, *AR* and *TERT*). The protective missense variant in *AR* is notable given the widespread treatment of prostate cancer patients with androgen deprivation therapy^3^, and highlights more broadly the utility of human population genetics for identifying potential therapeutic targets.

*SAMHD1* codes for an enzyme with diverse roles. It acts as a dNTPase, participates in innate immune regulation, maintains genome integrity and is recurrently somatically mutated across multiple tumour types^42,43^. It is notable that the same QV model strongly associated with prostate cancer risk was previously found to be associated with longer telomere length^44^. In combination with the confirmed association between genetically predicted longer telomere length and increased prostate cancer risk^38^, this implicates telomere maintenance as a potential mechanism for *SAMHD1*-mediated predisposition to prostate cancer. Conversely, rare deleterious variants in *TERT* are associated with shorter telomeres^44^, consistent with the finding here that a missense *TERT* variant is associated with a decreased risk of developing prostate cancer.

In our analysis, deleterious variants in *BRCA2* and *ATM* were associated with both the risk of developing prostate cancer and with a more aggressive form of the disease. The association of *BRCA2* and *ATM* with severity is in agreement with the statistically significant associations found in a recent large exome sequencing study of aggressive versus non-aggressive prostate cancer^45^. In comparison, two other genes – *SAMHD1* and *CHEK2* – showed significant associations in the case-control analysis of prostate cancer risk but demonstrated no association with severity. This suggests that they play a more prominent role in malignant transformation, with no substantial role in the progression of established cancer, similar to the reported effect of *HOXB13* p.Gly84Glu. Conversely, damaging germline variants in *AOX1* were significantly associated solely with aggressive forms of prostate cancer. This implies a role for *AOX1* in the progression of prostate cancer, specifically in determining severity, but no substantial impact on the overall risk of disease development.

The findings have potential clinical implications that warrant further investigation. Inheritance of variants associated with prostate cancer risk, for example, could influence prostate cancer screening recommendations, with carriers feasibly requiring earlier and/or more intensive testing. Inheritance of variants associated with aggressive prostate cancer could impact treatment decisions, with newly diagnosed carriers warranting more intensive treatment compared with non-carriers who share otherwise similar clinical characteristics. Furthermore, the identification of pathogenic variants in specific genes/pathways could inform precision medicine strategies. Finally, clinical risk stratification tools will likely be improved by integration of rare germline variants identified here with previously established risk factors, including common germline variants, somatic tumour driver mutations and non-genetic patient features.

Overall, our analysis provides novel insights into the contribution of rare deleterious variants to prostate cancer risk and severity and, through the associated genes, into pathogenic mechanisms. While some overlap exists, we observe different genetic determinants of risk and severity, with critical implications for potentially distinct therapeutic approaches to prostate cancer prevention and treatment, respectively.

## Methods

### Cohorts

We brought together data from prospective cohort studies of cancer-free men as well as clinical and epidemiologic studies of patients with prostate cancer. The UK Biobank (UKB) is a prospective study which recruited approximately 500,000 participants between the ages of 40 and 65 years in the United Kingdom from 2006 until 2010, of whom 46% were male^16^. Each participant provided blood and urine samples. Additionally, data for each patient includes periodically updated electronic health records, health questionnaire results, and linkage to death and cancer registries. The study has approval from the North-West Multi-centre Research Ethics Committee (11/NW/0382), and participants provided informed consent.

The Mexico City Prospective Study (MCPS) is a cohort of approximately 150,000 participants recruited at 35 years of age or older in Mexico City from 1998 to 2004, of whom 33% are male^19^. Participants provided a blood sample, completed a health questionnaire and the study provided access to their death registry data. The study was approved by the Mexican National Council for Science and Technology, the Mexican Ministry of Health and the University of Oxford ethics committees.

The New York-Boston-AstraZeneca (NYBAZ) prostate cancer study consists of prostate cancer patients from three separate cohorts: participants of the Health Professionals Follow-up Study (HPFS) and the Physicians’ Health Study (PHS) who were diagnosed with prostate cancer during prospective follow-up and patients with cancer seen at the Dana-Farber Cancer Institute (DFCI) Gelb Center. From these three studies, 2,607 participants with high-risk prostate cancer were selected who had blood samples available.

HPFS and PHS are prospective cohorts that enrolled men from across the US with a professional background in health professions (HPFS) and medicine (PHS). HPFS started in 1986 with 51,529 initially cancer-free men, collected blood samples in 1993– 95 from 18,000, and continues to follow participants for cancer incidence and mortality. PHS started as randomized-controlled trials of aspirin and multivitamins in chronic disease prevention among 22,071 initially cancer-free men in 1982, with blood samples at baseline. Follow-up for both cohorts is similar, and prostate cancer diagnoses were confirmed by a review of medical records and pathology reports.^46^ Causes of death were assigned by a physician endpoint committee based on medical records, reports from next-of-kin, and the National Death Index. Data for this study included those with a prostate cancer diagnosis (1982–2014) with an available blood sample, who were high-risk (Gleason score ≥4+3 (grade groups 3–5), stage ≥T3, or PSA ≥20 ng/ml), but no regional or distant metastases at diagnosis (cN0/Nx M0/Mx or pN0/Nx M0/Mx).

DFCI GELB is an observational clinical study that includes patients with prostate cancer seen in the medical oncology department since 1997. Demographic and clinical data were captured in a structured database by treating clinicians at enrolment or by research assistants from the electronic medical record during follow-up,^47^ with death follow-up via the National Death Index. Patients (1997–2018) were selected for sequencing if they had localized (N0 M0) prostate cancer at initial diagnosis, had undergone surgery or radiation, had at least one high-risk feature as in HPFS/PHS (except Gleason scores ≥8/grade group 4–5), had any repeat contact with DFCI (95%), and had survived ≥3 years after initial diagnosis.

The 100,000 Genomes Project recruited patients from the United Kingdom’s National Health Service based on rare disease and cancer diagnoses^20,21^. Blood samples and clinical data were collected, and with consent participants were linked to electronic health records and the UK cancer registry. This study was approved by the National Research Ethics Committee.

The AstraZeneca clinical trial cohort contained a total of 1,445 prostate cancer patients enrolled across nine clinical trials: EPOC (NCT00090363), ENTHUSE M1 (NCT00554229), ENTHUSE M0 (NCT00626548), ENTHUSE M1C (NCT00617669), UVA97934;Study 8 (NCT01972217), PROpel (NCT03732820), MAD (NCT04087174), NCT04089553, AARDVARC (NCT04495179). All enrolled patients were diagnosed with either metastatic prostate cancer, castration-resistant prostate cancer, or metastatic castration-resistant prostate cancer.

FinnGen is a research project encompassing 9 Finnish biobanks, and the results presented here are from approximately 445,000 participants included in FinnGen release 11^15^. Blood samples were collected from each participant and data from the Finnish nationwide longitudinal health register is available. Study approval was obtained by the Coordinating Ethics Committee of the Hospital District of Helsinki and Uusimaa (number HUS/990/2017).

### Phenotypes

In the UK Biobank, prostate cancer cases were identified from the cancer register (UKB Data-Field 40006), death register (UKB Data-Fields 40001 and 40002) and hospital inpatient diagnoses (UKB Data-Fields 41270) using International Classification of Diseases (ICD)-10 code C61, and additional cases from primary care records (Read v2). In the MCPS cohort cases were identified as participants self-reporting as diagnosed with prostate cancer in the baseline recruitment questionnaire and from the death register (ICD-10 code C61). In the 100,000 Genomes Project cohort cases were identified from those recruited to the project based on a diagnosis of prostate cancer. Additional cases were identified across the entire project cohort from linkage to the hospital episode statistics, the cancer register and the death register using ICD-10 code C61. All individuals in the AstraZeneca clinical trial cohort were recruited to the trials based on a diagnosis of prostate cancer. Finally, in the FinnGen cohort cases were identified from hospital discharge records, cause of death records and cancer registry using ICD-10 code C61 and ICD-9 code 185.

Controls in UK Biobank were used for the UKB and NYBAZ cohorts of cases. These were defined as male participants without malignant neoplasm diagnoses, as defined by ICD-10 codes C00-C90 in the cancer register, hospital admissions, and death register. Additionally, individuals were removed from the control set based on self-reported prostate cancer or family history of prostate cancer (father or brother diagnosed with prostate cancer). UKB controls for the NYBAZ cohort were selected based on those samples in the UKB which best matched the total number of rare deleterious variants across the exome (“flexdmg” QV model as in Supplementary Table 1). In MCPS, controls were defined as male participants without prostate cancer. Controls for the AstraZeneca clinical trials prostate cancer case cohort were comprised of male participants from non-oncology clinical trials in the cardiovascular, renal, metabolism, respiratory and immunology therapy areas. For the 100,000 Genomes Project, a set of controls was identified from the rare disease arm of the project. From these, male individuals who were not the proband and who had no prostate cancer diagnosis were selected. In the FinnGen study, male participants with no diagnoses of any cancer were used as controls.

In the UKB, MCPS, NYBAZ, 100,000 Genomes Project and AZ clinical trials cohorts, cases were stratified into non-aggressive prostate cancer and aggressive prostate cancer based on the available clinical data. In the UKB and 100,000 Genomes Project, aggressive prostate cancer cases were defined as those with prostate cancer as the underlying cause of death or prostate cancer as the only primary neoplasm and a secondary neoplasm (ICD-10 codes C77, C78, C79) or prostate cancer and chemotherapy (based on OPCS Classification of Interventions and Procedures). In MCPS, aggressive prostate cancer was identified as those with prostate cancer as their underlying cause of death. In the NYBAZ cohort, individuals with tumour stage T4/N1 or Gleason score >=8 were defined as aggressive prostate cancer. All participants in the AstraZeneca clinical trials cohort were metastatic and/or castration-resistant and were therefore classified as aggressive prostate cancer cases.

### Sequencing, Variant Calling, Genotyping and Imputation

For all whole exome sequencing studies sequencing was performed using the IDT xGen v1 capture kit on the NovaSeq6000 platform. Both the UKB and MCPS cohorts were whole exome sequenced at the Regeneron Genetics Center with 75-bp paired ends^18,23,48^. The New York-Boston-AstraZeneca (NYBAZ) prostate cancer study samples were whole exome sequenced at the Institute for Genomic Medicine at the Columbia University Medical Center with 150-bp paired ends. All AstraZeneca clinal trial whole exome sequencing was performed at Human Longevity Inc. with 150-bp paired-ends.

All whole exome sequencing FASTQ data was processed at AstraZeneca using Amazon Web Services cloud computing platform. Reads were aligned to the GRCh38 genome reference, and small variant calling performed, with the Illumina DRAGEN Bio-IT Platform Germline Pipeline v3.0.7. Variants were annotated with SnpEFF v4.3^49^ against Ensembl Build 38.92 and with their genome Aggregation Database (gnomAD) MAFs (gnomAD v2.1.1 mapped to GRCh38)^50^.

As previously described^20^, the 100,000 Genomes Project was whole genome sequenced using TruSeq DNA polymerase-chain-reaction (PCR)–free sample preparation kit (Illumina) on the HiSeq2500 platform. Reads were aligned using the Isaac Genome Alignment Software, and small variant calling performed with the Platypus variant caller^51^. Variants were annotated with VEP v105 with the gnomAD plugin included^52^.

FinnGen genotyping and imputation has been previously described^15^. In brief, genotyping was performed with Illumina (Illumina) and Affymetrix arrays (Thermo Fisher Scientific) and calls with GenCall and zCall algorithms. Imputation was carried out using Beagle 4.1 with a reference panel generated from the whole genome sequencing of 3,775 Finnish individuals.

### Cohort Harmonisation and Quality Control

All whole exome sequenced cohorts underwent quality control as previously described^23,53^. Pre-harmonisation and quality control the UKB cohort consisted of 15,417 cases and 147,652 male controls; the MCPS cohort of 287 cases and 46,717 male controls; the NYBAZ cohort of 2,506 cases; the clinical trials cohort of 1,445 cases. In brief, samples were excluded if contaminated (VerifyBamID contamination >= 4%), and if there was discordance between the self-reported and genetically determined sex. Samples were only included for downstream analysis if they achieved ≥94.5% of consensus coding sequence (CCDS) r22 bases covered with ≥10-fold coverage. We excluded participants that were second-degree relatives or closer, estimated with KING v2.2.3^54^ using the --kinship function (kinship coefficient > 0.0884). Continent level ancestry was predicted using PEDDY v0.4.2^55^ with the 1000 Genomes Project sequences as a population reference. For European cohorts, only individuals with a predicted probability greater than 99% of European ancestry were selected. Non-European strata were included if there were a minimum of 75 cases and the probability threshold was set at greater than 95% for the relevant ancestry. Additionally, only individuals who were within 4 SD of the cohort means for the top four principal components were selected. Finally, samples outside 4 SD of the mean for novel CCDS SNPs in the test cohort were excluded.

For the 100,000 Genomes Project whole genome sequenced cohort, a similar set of harmonisation steps were performed. Before harmonisation the cohort consisted of 1,347 cases and 32,985 controls. Pre-harmonisation QC was performed on all whole genome sequences: samples were required not to be contaminated (VerifyBamID freemix <= 3%); aligned reads were required to cover 95% of the genome at 15X or above with mapping quality >10; array concordance >90%; median fragment size > 250bp; chimeric reads < 5%; median fragment size > 250bp; mapped reads > 60%; AT dropout < 10%; self-reported and genetically determined sex were required to match. For cohort harmonisation, continental ancestry was predicted by training a random forest model on eight 1KGP3 PCs, and only individuals with a probability of European ancestry greater than 99% were selected. Additionally, only individuals who were within 4 SD of the cohort means for the top four principal components were selected. Finally, participants that were second-degree relatives or closer were removed (prioritising retaining cases), as estimated with KING.

Sample quality control for the FinnGen cohort was as previously described^15^, and consisted of ensuring genetically determined sex matched reported sex, low genotype missingness (<5%) and low heterozygosity (±4 standard deviations).

### Gene-level Collapsing Analysis

As previously described, we performed gene-level collapsing analysis across eleven QV models^23^, (Supplementary Table 1). For dominant collapsing models, carriers with at least one qualifying variant were tested against non-carriers. For the single recessive QV model, carriers were defined as those with a homozygous qualifying variant, or at least two heterozygous qualifying variants (i.e. putatively compound heterozygous). The association of qualifying variant carriers with prostate cancer risk and its severity was tested for with Fisher’s exact two-sided test within each cohort. Meta-analysis across cohorts was performed with the Cochran–Mantel–Haenszel (CMH) test. We excluded 56 genes that we previously found to be associated with batch effects. To DRAGEN whole exome sequencing PASS variant calls we applied additional filters: coverage >= 10x; CCDS transcripts annotation (release 22); heterozygous variant alternative allele reads >= 0.3 and <= 0.8; alternate allele percentage significantly different from 50% in heterozygous state (binomial P > 1 × 10−6); read position rank sum score (RPRS) ≥ −2; genotype quality score (GQ) ≥ 30; Fisher’s strand bias score (FS) ≤ 200 (indels) ≤ 60 (SNVs); quality score (QUAL) ≥ 30; mapping quality score (MQ) ≥ 40; mapping quality rank sum score (MQRS) ≥ −8; in ≥25% of gnomAD exomes the site achieved 10-fold coverage; if in gnomAD exomes the variant was observed then we required exome z score ≥ −2.0 and exome MQ ≥ 30.

### Exome Wide Association Analysis

For next generation sequenced cohorts, single-variant association testing for exome variants was performed as previously described^23^. Variant association with prostate cancer risk and its severity was tested for with Fisher’s exact two-sided test under three genetic models: dominant (XX + XY versus YY), allelic (X versus Y) and recessive (XX versus XY + YY), where X is the alternate allele and Y is the ref allele. We applied to DRAGEN whole exome sequencing PASS variant calls additional filters: coverage >= 10×; homozygous variant alternative allele reads >= 0.8; heterozygous variant alternative allele reads >= 0.3 and <= 0.8; alternate allele percentage significantly different from 50% in heterozygous state (binomial P > 1 × 10^−6^); FS ≤ 200 (indels) ≤ 60 (SNVs) ; MQ ≥ 40; RPRS ≥ −2; QUAL ≥ 30; GQ ≥ 30; MQRS ≥ −8; the variant site does not have less than 10× coverage in 1% or more of sequences; the variant must not have failed any of these QC metrics in >0.5% sequences; in >50% of gnomAD exomes the variant site achieved >10x coverage; Hardy-Weinberg equilibrium test *P* < 1×10^-10^. Single variant association statistics for the risk of developing prostate cancer in the FinnGen cohort were generated with REGENIE^56^ under the additive model with sex, age and 10 principal components included as covariates (Pipeline details: https://github.com/FINNGEN/regenie-pipelines). Meta-analysis across sequenced cohorts for the dominant and recessive models was performed with CMH. Across all cohorts, meta-analysis was performed with the sample sized based (Stouffer’s) method as implemented in METAL^57^ using allelic or additive summary statistics as available.

## Data Availability

Summary statistics for the genetic associaiton testing are provided in Supplementary Tables. UK Biobank data may be requested via application to the UK Biobank.

## Acknowledgments

This research has been conducted using the UK Biobank Resource under Application Number 26041.

This research was made possible through access to data in the National Genomic Research Library, which is managed by Genomics England Limited (a wholly owned company of the Department of Health and Social Care). The National Genomic Research Library holds data provided by patients and collected by the NHS as part of their care and data collected as part of their participation in research. The National Genomic Research Library is funded by the National Institute for Health Research and NHS England. The Wellcome Trust, Cancer Research UK and the Medical Research Council have also funded research infrastructure.

## References

1. Cancer (IARC), T. I. A. for R. on. Global Cancer Observatory. https://gco.iarc.fr/.

2. Sung, H. et al. Global Cancer Statistics 2020: GLOBOCAN Estimates of Incidence and Mortality Worldwide for 36 Cancers in 185 Countries. CA. Cancer J. Clin. 71, 209–249 (2021).

3. Rebello, R. J. et al. Prostate cancer. Nat. Rev. Dis. Primer 7, 9 (2021).

4. Siegel, R. L., Miller, K. D. & Jemal, A. Cancer statistics, 2018. CA. Cancer J. Clin. 68, 7– 30 (2018).

5. Mucci, L. A. et al. Familial Risk and Heritability of Cancer Among Twins in Nordic Countries. JAMA 315, 68–76 (2016).

6. Conti, D. V. et al. Trans-ancestry genome-wide association meta-analysis of prostate cancer identifies new susceptibility loci and informs genetic risk prediction. Nat. Genet. 53, 65–75 (2021).

7. Schumacher, F. R. et al. Association analyses of more than 140,000 men identify 63 new prostate cancer susceptibility loci. Nat. Genet. 50, 928–936 (2018).

8. Wang, A. et al. Characterizing prostate cancer risk through multi-ancestry genome-wide discovery of 187 novel risk variants. Nat. Genet. 55, 2065–2074 (2023).

9. Zhiyu Yang et al. Limited overlap between genetic effects on disease susceptibility and disease survival. medRxiv 2023.10.10.23296544 (2023) doi:10.1101/2023.10.10.23296544.

10. Claussnitzer, M. et al. A brief history of human disease genetics. Nature 577, 179– 189 (2020).

11. de Bono, J. et al. Olaparib for Metastatic Castration-Resistant Prostate Cancer. N. Engl. J. Med. 382, 2091–2102 (2020).

12. Cohen, J. C., Boerwinkle, E., Mosley, T. H. & Hobbs, H. H. Sequence variations in PCSK9, low LDL, and protection against coronary heart disease. N. Engl. J. Med. 354, 1264–1272 (2006).

13. Ewing, C. M. et al. Germline mutations in HOXB13 and prostate-cancer risk. N. Engl. J. Med. 366, 141–149 (2012).

14. Pritchard, C. C. et al. Inherited DNA-Repair Gene Mutations in Men with Metastatic Prostate Cancer. N. Engl. J. Med. 375, 443–453 (2016).

15. Kurki, M. I. et al. FinnGen provides genetic insights from a well-phenotyped isolated population. Nature 613, 508–518 (2023).

16. Bycroft, C. et al. The UK Biobank resource with deep phenotyping and genomic data. Nature 562, 203–209 (2018).

17. Van Hout, C. V. et al. Exome sequencing and characterization of 49,960 individuals in the UK Biobank. Nature 586, 749–756 (2020).

18. Ziyatdinov, A. et al. Genotyping, sequencing and analysis of 140,000 adults from Mexico City. Nature 622, 784–793 (2023).

19. Tapia-Conyer, R. et al. Cohort profile: the Mexico City Prospective Study. Int. J. Epidemiol. 35, 243–249 (2006).

20. 100,000 Genomes Project Pilot Investigators et al. 100,000 Genomes Pilot on Rare-Disease Diagnosis in Health Care - Preliminary Report. N. Engl. J. Med. 385, 1868–1880 (2021).

21. Sosinsky, A. et al. Insights for precision oncology from the integration of genomic and clinical data of 13,880 tumors from the 100,000 Genomes Cancer Programme. Nat. Med. 30, 279–289 (2024).

22. Petrovski, S. et al. An Exome Sequencing Study to Assess the Role of Rare Genetic Variation in Pulmonary Fibrosis. Am. J. Respir. Crit. Care Med. 196, 82–93 (2017).

23. Wang, Q. et al. Rare variant contribution to human disease in 281,104 UK Biobank exomes. Nature 597, 527–532 (2021).

24. Stasa Stankovic et al. Genetic susceptibility to earlier ovarian ageing increases de novo mutation rate in offspring. medRxiv 2022.06.23.22276698 (2022) doi:10.1101/2022.06.23.22276698.

25. Wilcox, N. et al. Exome sequencing identifies breast cancer susceptibility genes and defines the contribution of coding variants to breast cancer risk. Nat. Genet. 55, 1435– 1439 (2023).

26. Cavazos, T. B. et al. Assessment of genetic susceptibility to multiple primary cancers through whole-exome sequencing in two large multi-ancestry studies. BMC Med. 20, 332 (2022).

27. AstraZeneca PheWAS Portal. https://azphewas.com/geneView/ba08a93f-501e-44e6-a332-98ce2f852279/SAMHD1/glr/binary.

28. Jones, L., Naidoo, M., Machado, L. R. & Anthony, K. The Duchenne muscular dystrophy gene and cancer. Cell. Oncol. Dordr. 44, 19–32 (2021).

29. Kar, S. P. et al. Genome-wide analyses of 200,453 individuals yield new insights into the causes and consequences of clonal hematopoiesis. Nat. Genet. 54, 1155–1166 (2022).

30. Li, W. et al. Genome-wide Scan Identifies Role for AOX1 in Prostate Cancer Survival. Eur. Urol. 74, 710–719 (2018).

31. Wang, G., Sarkar, A., Carbonetto, P. & Stephens, M. A simple new approach to variable selection in regression, with application to genetic fine mapping. J. R. Stat. Soc. Ser. B Stat. Methodol. 82, 1273–1300 (2020).

32. Nyberg, T. et al. Prostate Cancer Risks for Male BRCA1 and BRCA2 Mutation Carriers: A Prospective Cohort Study. Eur. Urol. 77, 24–35 (2020).

33. Kote-Jarai, Z. et al. Prevalence of the HOXB13 G84E germline mutation in British men and correlation with prostate cancer risk, tumour characteristics and clinical outcomes. Ann. Oncol. Off. J. Eur. Soc. Med. Oncol. 26, 756–761 (2015).

34. Kaikkonen, E. et al. ANO7 is associated with aggressive prostate cancer. Int. J. Cancer 143, 2479–2487 (2018).

35. Barisic, M. et al. Spindly/CCDC99 is required for efficient chromosome congression and mitotic checkpoint regulation. Mol. Biol. Cell 21, 1968–1981 (2010).

36. Dhindsa, R. S. et al. Identification of a missense variant in SPDL1 associated with idiopathic pulmonary fibrosis. *Commun*. Biol. 4, 392 (2021).

37. Vancheri, C. Common pathways in idiopathic pulmonary fibrosis and cancer. Eur. Respir. Rev. Off. J. Eur. Respir. Soc. 22, 265–272 (2013).

38. Codd, V. et al. Polygenic basis and biomedical consequences of telomere length variation. Nat. Genet. 53, 1425–1433 (2021).

39. Rode, L., Nordestgaard, B. G. & Bojesen, S. E. Long telomeres and cancer risk among 95 568 individuals from the general population. Int. J. Epidemiol. 45, 1634–1643 (2016).

40. DeBoy, E. A. et al. Familial Clonal Hematopoiesis in a Long Telomere Syndrome. N. Engl. J. Med. 388, 2422–2433 (2023).

41. Berchuck, J. E. et al. Addition of Germline Testing to Tumor-Only Sequencing Improves Detection of Pathogenic Germline Variants in Men With Advanced Prostate Cancer. *JCO Precis*. Oncol. 6, e2200329 (2022).

42. Coggins, S. A., Mahboubi, B., Schinazi, R. F. & Kim, B. SAMHD1 Functions and Human Diseases. Viruses 12, 382 (2020).

43. Schott, K. et al. SAMHD1 in cancer: curse or cure? J. Mol. Med. Berl. Ger. 100, 351– 372 (2022).

44. Oliver S. Burren et al. Genetic architecture of telomere length in 462,675 UK Biobank whole-genome sequences. medRxiv 2023.09.18.23295715 (2023) doi:10.1101/2023.09.18.23295715.

45. Darst, B. F. et al. Germline Sequencing Analysis to Inform Clinical Gene Panel Testing for Aggressive Prostate Cancer. JAMA Oncol. 9, 1514–1524 (2023).

46. Giovannucci, E., Liu, Y., Platz, E. A., Stampfer, M. J. & Willett, W. C. Risk factors for prostate cancer incidence and progression in the health professionals follow-up study. Int J Cancer 121, 1571–8 (2007).

47. Oh, W. K. et al. Development of an integrated prostate cancer research information system. Clin Genitourin Cancer 5, 61–6 (2006).

48. Backman, J. D. et al. Exome sequencing and analysis of 454,787 UK Biobank participants. Nature 599, 628–634 (2021).

49. Cingolani, P. et al. A program for annotating and predicting the effects of single nucleotide polymorphisms, SnpEff: SNPs in the genome of Drosophila melanogaster strain w1118; iso-2; iso-3. Fly (Austin) 6, 80–92 (2012).

50. Karczewski, K. J. et al. The mutational constraint spectrum quantified from variation in 141,456 humans. Nature 581, 434–443 (2020).

51. Rimmer, A. et al. Integrating mapping-, assembly- and haplotype-based approaches for calling variants in clinical sequencing applications. Nat. Genet. 46, 912–918 (2014).

52. McLaren, W. et al. The Ensembl Variant Effect Predictor. Genome Biol. 17, 122 (2016).

53. Nag, A. et al. Human genetics uncovers MAP3K15 as an obesity-independent therapeutic target for diabetes. Sci. Adv. 8, eadd5430 (2022).

54. Manichaikul, A. et al. Robust relationship inference in genome-wide association studies. Bioinforma. Oxf. Engl. 26, 2867–2873 (2010).

55. Pedersen, B. S. & Quinlan, A. R. Who’s Who? Detecting and Resolving Sample Anomalies in Human DNA Sequencing Studies with Peddy. Am. J. Hum. Genet. 100, 406– 413 (2017).

56. Mbatchou, J. et al. Computationally efficient whole-genome regression for quantitative and binary traits. Nat. Genet. 53, 1097–1103 (2021).

57. Willer, C. J., Li, Y. & Abecasis, G. R. METAL: fast and efficient meta-analysis of genomewide association scans. Bioinforma. Oxf. Engl. 26, 2190–2191 (2010).

