## Supplementary Figures for "Characterising the contribution of rare protein-coding germline variants to prostate cancer risk and severity in 37,184 cases"

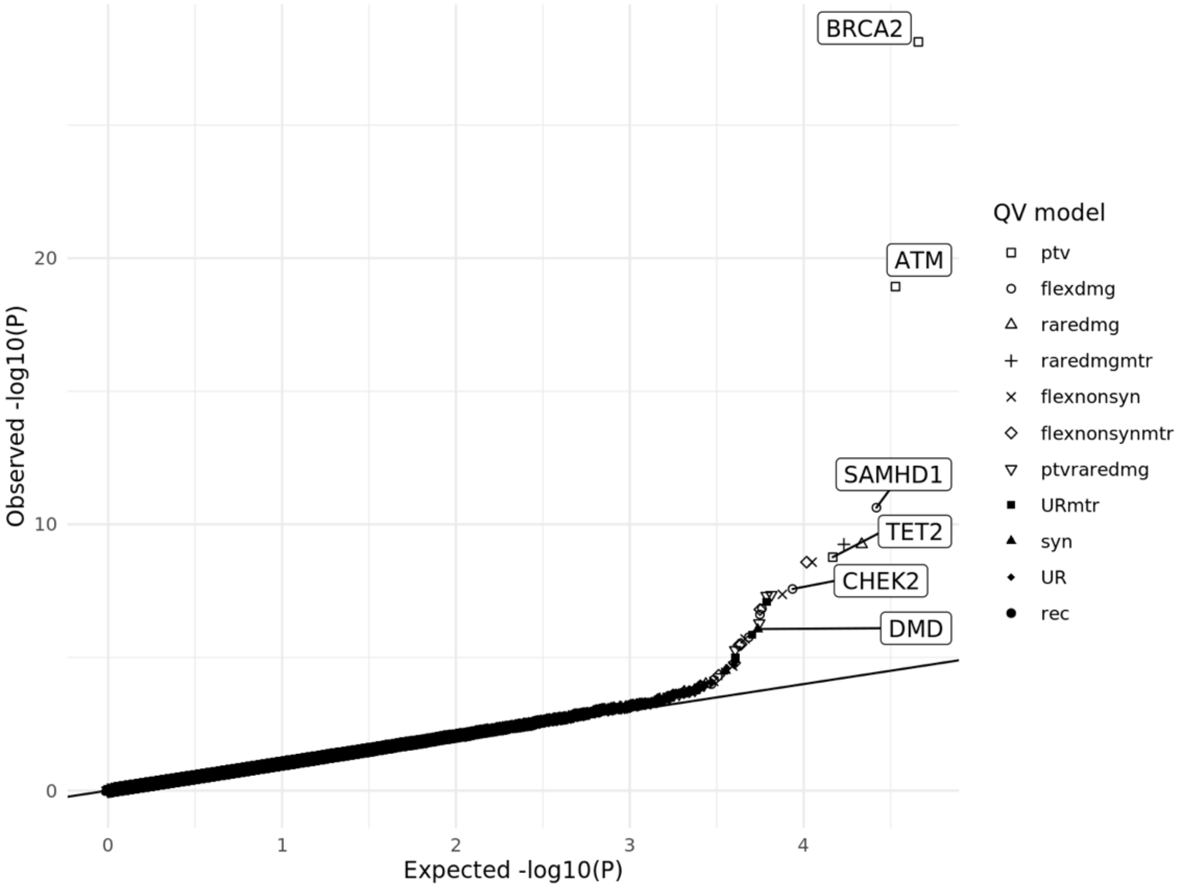


**Supplementary Figure 1: QQ-plot for meta-analysis of gene-level association tests for all qualifying variant models for risk of developing prostate cancer (cases versus controls).** Expected *P*-values on x-axis are generated from n-1 case-control permutation. Genes which reach the suggestive significance threshold ($P < 2.6 \times 10^{-6}$) are labelled, and only the most significant qualifying variant model for each gene is labelled.

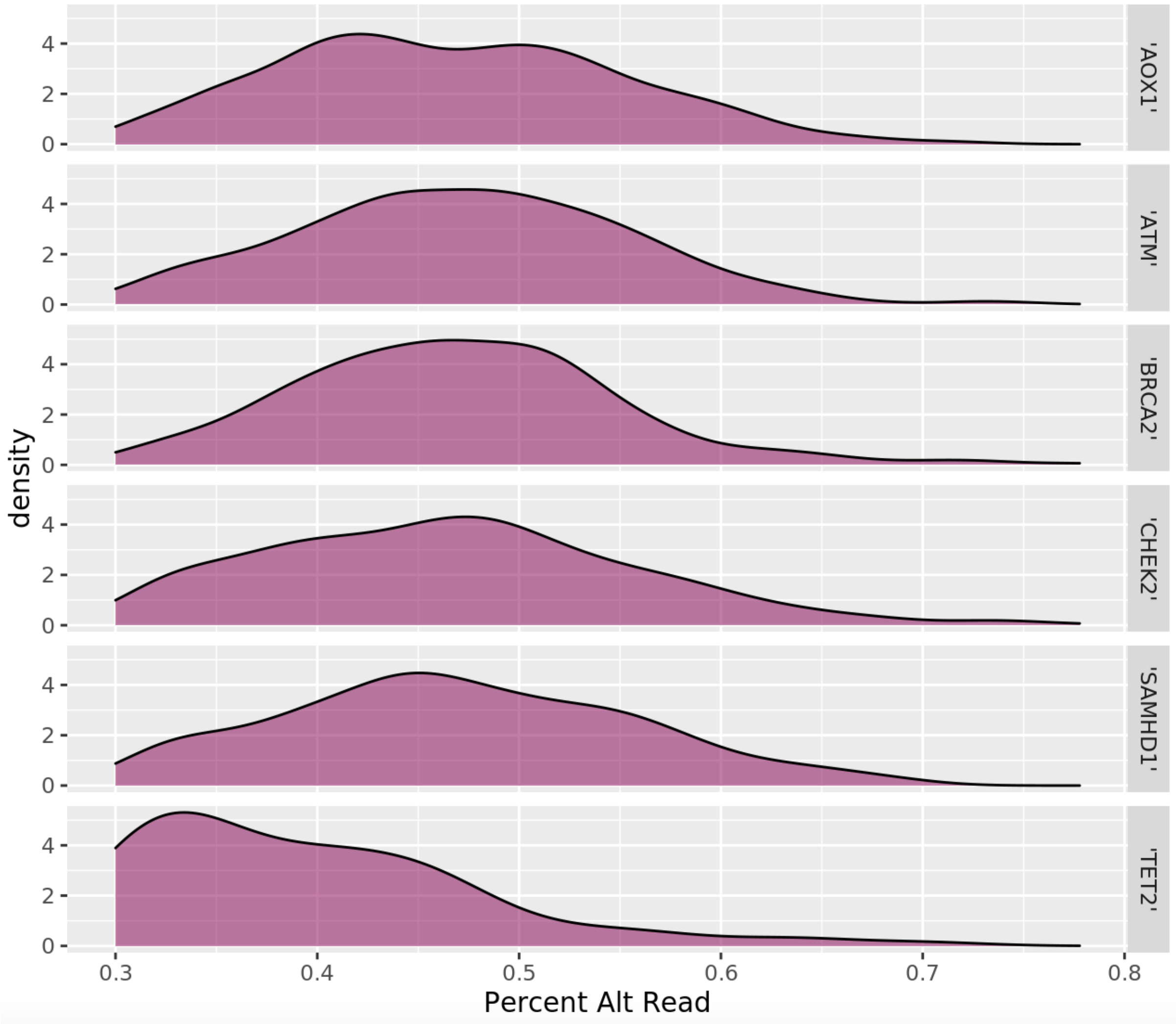


**Supplementary Figure 2: Distribution of alternative reads percentage for qualifying variants in genes associated with prostate cancer.**

| Gene | Odds Ratio (95% CI) | P-value |
| --- | --- | --- |
| BRCA2 (ptv) | 3.23 (2.65-3.90) | 7.5e-29 |
| ATM (ptv) | 2.92 (2.34-3.63) | 1.17e-19 |
| CHEK2 (flexdmg) | 1.69 (1.41-2.01) | 2.69e-08 |
| MSH2 (ptv) | 3.38 (1.55-6.90) | 0.0012 |
| NBN (raredmgmtr) | 1.92 (1.22-2.93) | 0.00335 |
| BRIP1 (raredmgmtr) | 0.68 (0.46-0.97) | 0.0293 |
| PALB2 (URmtr) | 1.75 (1000-2.91) | 0.0415 |
| RAD51C (flexnonsynmtr) | 1.38 (0.96-1.94) | 0.0656 |
| XRCC2 (ptv) | 1.68 (0.91-2.94) | 0.0659 |
| MSH6 (flexnonsynmtr) | 1.15 (0.99-1.34) | 0.0693 |
| MLH1 (ptv) | 2.03 (0.86-4.29) | 0.0882 |
| GEN1 (UR) | 1.55 (0.87-2.60) | 0.0948 |
| BAP1 (ptv) | 2.50 (0.58-8.38) | 0.113 |
| RAD51D (raredmg) | 0.64 (0.35-1.08) | 0.114 |
| PMS2 (syn) | 1.27 (0.93-1.71) | 0.121 |
| ATR (flexdmg) | 1.10 (0.94-1.30) | 0.223 |
| BRCA1 (URmtr) | 0.80 (0.55-1.14) | 0.255 |
| BARD1 (syn) | 1.19 (0.85-1.62) | 0.278 |

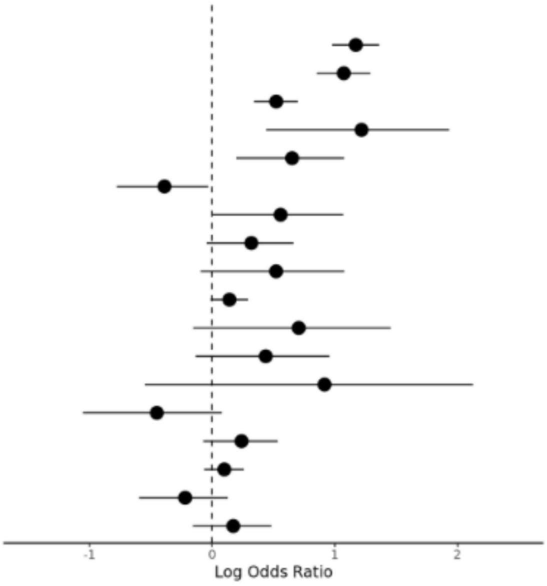


**Supplementary Figure 3: Forest plots showing the association of DNA damage response genes with the development of prostate cancer (cases versus controls).** Results are from the gene-level collapsing analysis, and only the qualifying variant model with the lowest *P*-value for each gene is displayed (as indicated in column one parentheses).

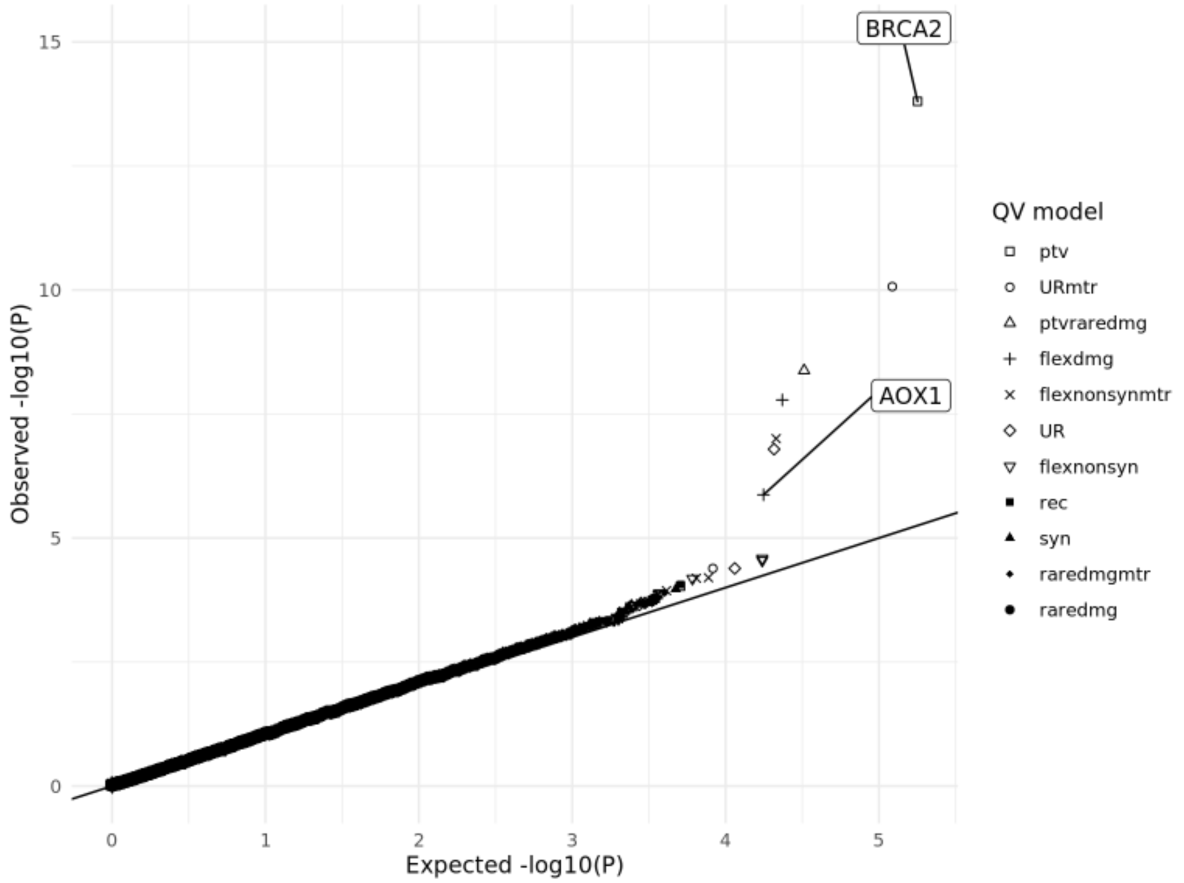


**Supplementary Figure 4: QQ-plot for meta-analysis of gene-level association tests for all qualifying variant models for prostate cancer severity (aggressive prostate cancer vs non- aggressive prostate cancer).** Expected *P*-values on x-axis are generated from n-1 case-control permutation. Genes which reach the suggestive significance threshold ($P < 2.6\text{x}10^{-6}$) are labelled, and only the most significant qualifying variant model for each gene is labelled.

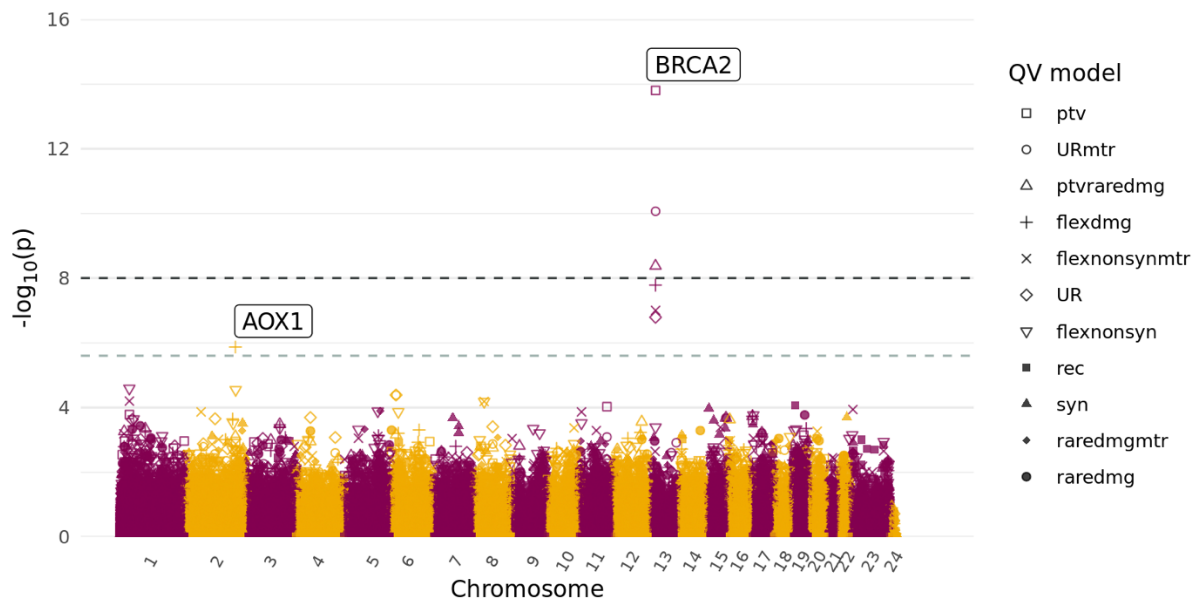


**Supplementary Figure 5: Manhattan plot of all gene-level association tests with aggressive prostate cancer versus non-aggressive prostate cancer.** The x-axis is the genomic position of the gene, and the y-axis is the $-\log_{10}$ transformed unadjusted *P*-values for all qualifying variant models as indicated in the legend. The light grey dashed line represents the suggestive significance threshold ($P = 2.6\text{x}10^{-6}$) and the dark grey dashed line the study-wide significance threshold ($P = 1\text{x}10^{-8}$). Genes which reach the suggestive significance threshold are labelled, and only the most significant qualifying variant model for each gene is labelled.

| Gene | Odds Ratio (95% CI) | P-value |
|---|---|---|
| BRCA2 (ptv) | 3.82 (2.70-5.41) | 1.58e-14 |
| ATM (ptv) | 2.23 (1.47-3.34) | 9.41e-05 |
| BARD1 (flexnonsynmtr) | 1.76 (1.12-2.72) | 0.0103 |
| MSH6 (raredmg) | 1.51 (1.07-2.10) | 0.0143 |
| BRCA1 (raredmg) | 1.49 (1.05-2.09) | 0.0217 |
| GEN1 (ptv) | 2.65 (0.98-6.71) | 0.0314 |
| RAD51C (URmtr) | 6.76 (0.77-80.82) | 0.0448 |
| RAD51D (syn) | 2.16 (0.88-5000) | 0.0631 |
| PALB2 (ptv) | 1.96 (0.91-4.02) | 0.0663 |
| MLH1 (ptv) | 3.75 (0.69-18.65) | 0.0675 |
| NBN (flexnonsyn) | 1.36 (0.93-1.97) | 0.0945 |
| MSH2 (ptv) | 2.54 (0.60-9.70) | 0.149 |
| CHEK2 (UR) | 1.80 (0.75-4.05) | 0.175 |
| BRIP1 (raredmgmtr) | 1.67 (0.69-3.78) | 0.189 |
| XRCC2 (ptv) | 0.24 (0.01-1.70) | 0.2 |
| ATR (syn) | 1.33 (0.80-2.15) | 0.251 |
| PMS2 (flexnonsynmtr) | 0.76 (0.40-1.34) | 0.359 |
| BAP1 (syn) | 1.29 (0.65-2.46) | 0.406 |

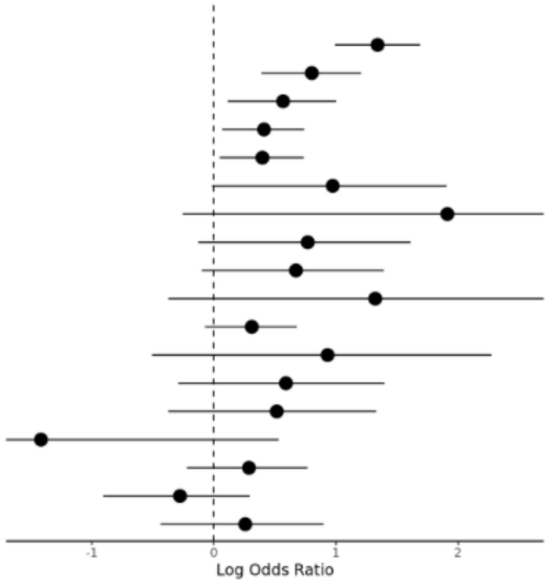


**Supplementary Figure 6: Forest plots showing the association of DNA damage response genes with aggressive prostate cancer versus non-aggressive prostate cancer**. Results are from the gene-level collapsing analysis, and only the qualifying variant model with the lowest *P*-value for each gene is displayed (as indicated in column one parentheses).

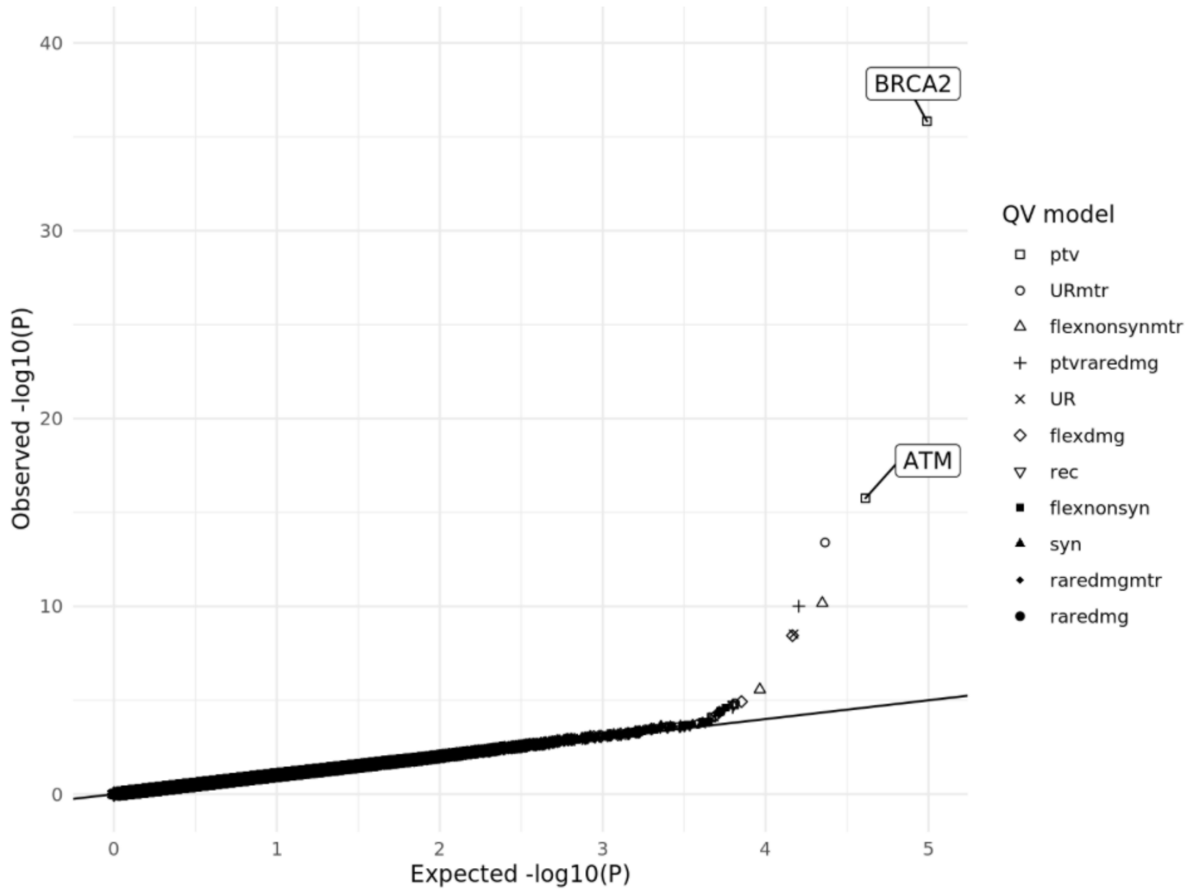


**Supplementary Figure 7: QQ-plot of all meta-analyses gene-level association tests with aggressive prostate cancer versus controls.** Expected *P*-values on x-axis are generated from n-1 case-control permutation. Genes which reach the suggestive significance threshold are labelled, and only the most significant qualifying variant model for each gene is labelled.

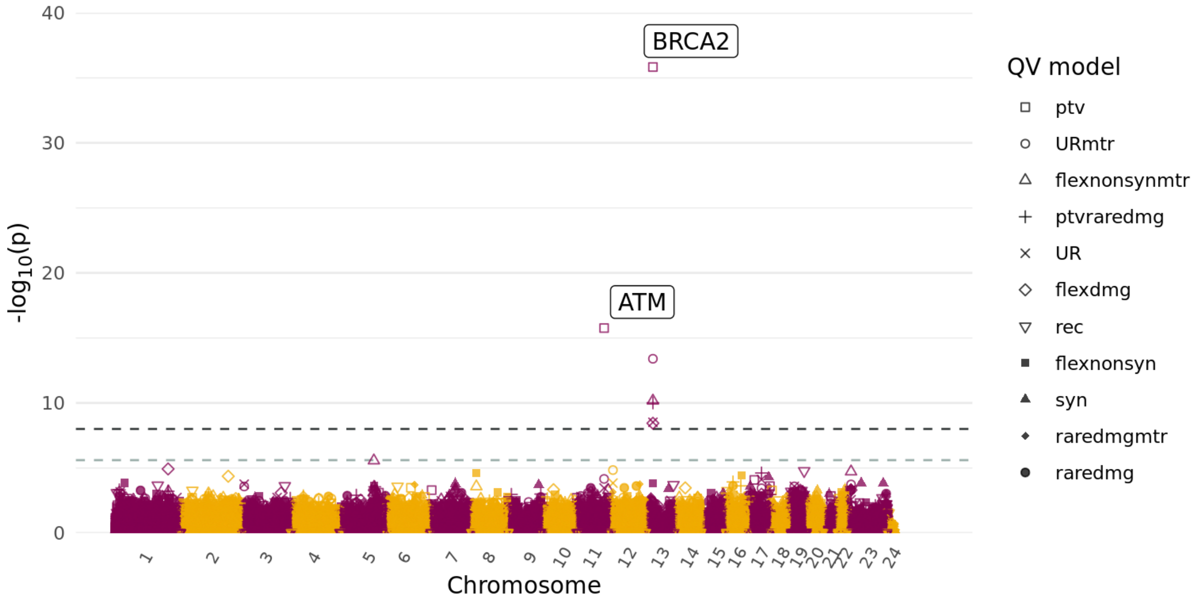


**Supplementary Figure 8: Manhattan plot of all gene-level association tests with aggressive prostate cancer versus controls.** The x-axis is the genomic position of the gene, and the y-axis is the $-\log_{10}$ transformed unadjusted *P*-values for all qualifying variant models as indicated in the legend. The light grey dashed line represents the suggestive significance threshold ($P = 2.6\text{x}10^{-6}$) and the dark grey dashed line the study-wide significance threshold ($P = 1\text{x}10^{-8}$). Genes which reach the suggestive significance threshold are labelled, and only the most significant qualifying variant model for each gene is labelled.

| Gene | Odds Ratio (95% CI) | P-value |
|---|---|---|
| BRCA2 (ptv) | 8.23 (6.17-10.85) | 1.47e-36 |
| ATM (ptv) | 5.27 (3.65-7.46) | 1.74e-16 |
| CHEK2 (flexnonsyn) | 1.67 (1.24-2.22) | 0.000702 |
| MSH2 (ptv) | 7.70 (1.99-23.31) | 0.00198 |
| MLH1 (ptv) | 5.89 (1.49-16.8) | 0.00678 |
| BRCA1 (raredmg) | 1.43 (1.08-1.88) | 0.0112 |
| RAD51D (syn) | 2.37 (1.13-4.48) | 0.0166 |
| NBN (raredmgmtr) | 2.64 (1.04-5.77) | 0.0203 |
| PALB2 (ptv) | 2.18 (1.09-4.01) | 0.0206 |
| MSH6 (ptvraredmg) | 1.33 (1000-1.74) | 0.0447 |
| GEN1 (URmtr) | 2.50 (0.82-6.24) | 0.053 |
| BARD1 (syn) | 1.69 (0.91-2.94) | 0.0641 |
| BAP1 (syn) | 1.69 (0.91-2.91) | 0.0832 |
| BRIP1 (flexnonsynmtr) | 1.33 (0.89-1.93) | 0.135 |
| RAD51C (flexnonsynmtr) | 1.59 (0.74-3.06) | 0.166 |
| ATR (syn) | 1.27 (0.85-1.85) | 0.213 |
| PMS2 (syn) | 1.37 (0.70-2.47) | 0.306 |
| XRCC2 (flexnonsynmtr) | 1.27 (0.65-2.26) | 0.414 |

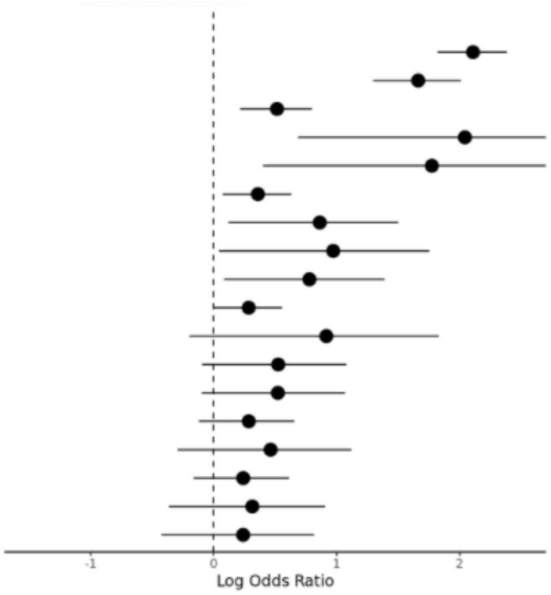


**Supplementary Figure 9: Forest plots showing the association of DNA damage response genes with aggressive prostate cancer versus controls**. Results are from the gene-level collapsing analysis, and only the qualifying variant model with the lowest *P*-value for each gene is displayed (as indicated in column one parentheses).

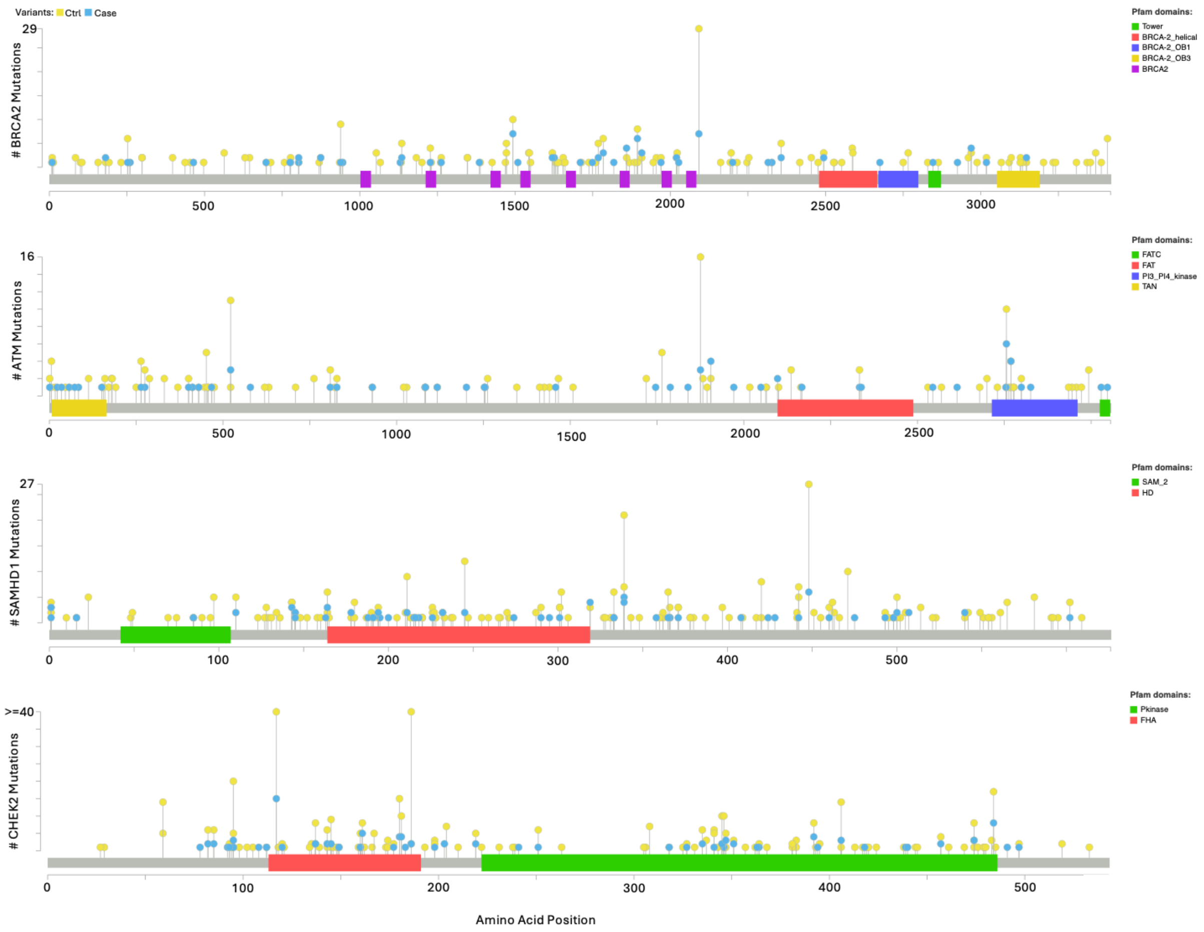


**Supplementary Figure 10: Lollipop plots showing the number of qualifying variants (y-axis) for genes significantly associated with the risk of developing prostate cancer (cases versus controls).** For each gene, only the most significantly associated qualifying variant model is shown (*BRCA2* = “ptv”; *ATM* = “ptv”; *SAMHD1* = “flexdmg”; *CHEK2* = “flexdmg”). Variants are from UK Biobank European carriers.

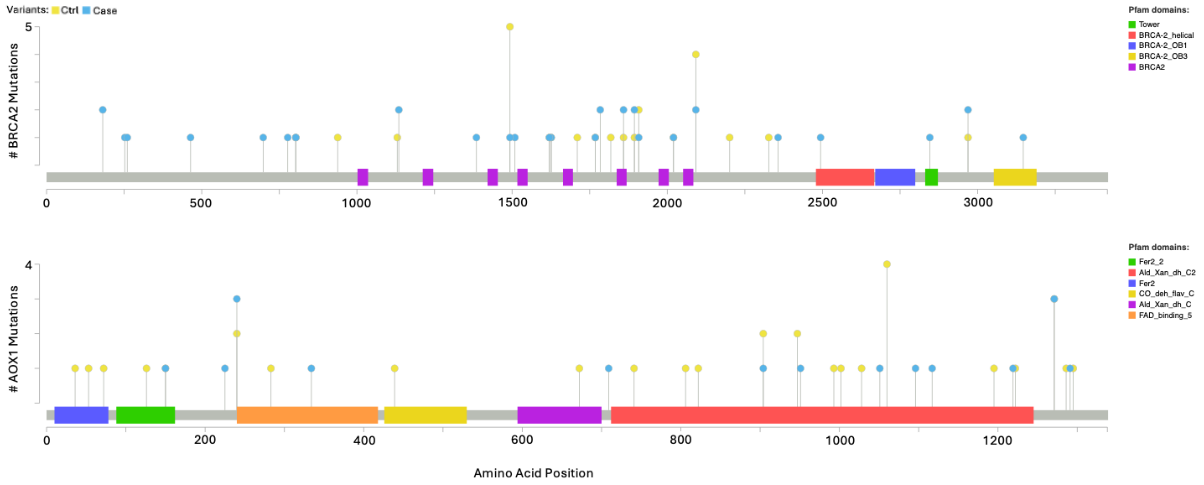


**Supplementary Figure 11: Lollipop plots showing the number of qualifying variants (y-axis) for genes significantly associated with aggressive prostate cancer versus non-aggressive prostate cancer.** For each gene, only the most significantly associated qualifying variant model is shown (*BRCA2* = "ptv"; *AOX1* = "flexdmg"). Variants are from UK Biobank European carriers. ctrl = non-aggressive prostate cancer, case = aggressive prostate cancer.
